# Psychosocial adversity experienced in utero and early life is associated with variation in gut microbiota: a prospective case-control study

**DOI:** 10.1101/2022.11.17.22282482

**Authors:** Barbara B. Warner, Bruce A. Rosa, I. Malick Ndao, Phillip I. Tarr, J. Phillip Miller, Sara K. England, Joan L. Luby, Cynthia E. Rogers, Carla Hall-Moore, Renay E. Bryant, Jacqueline D. Wang, Laura Linneman, Tara A. Smyser, Christopher D. Smyser, Deanna M. Barch, Gregory E. Miller, Edith Chen, John Martin, Makedonka Mitreva

**Affiliations:** Department of Pediatrics, Washington University School of Medicine, St. Louis, MO 63110; Department of Medicine, Washington University School of Medicine, St. Louis, MO 63110; Department of Molecular Microbiology, Washington University School of Medicine, St. Louis, MO 63110; Division of Biostatistics, Office of Health Information and Data Science, Washington University School of Medicine in St. Louis, St. Louis, MO 63110; Department of Obstetrics and Gynecology, Washington University in St. Louis, St. Louis, Missouri, 63110; Department of Psychiatry, Washington University School of Medicine in St. Louis, St. Louis, MO 63110; Departments of Neurology, Pediatrics and Radiology, Washington University School of Medicine in St. Louis, MO 63110; Department of Psychological and Brain Sciences, Washington University in St. Louis, St. Louis, MO 63130; Institute for Policy Research & Department of Psychology, Northwestern University, Evanston, Illinois 60208; Departments of Medicine, Genetics and McDonnell Genome Institute, Washington University School of Medicine in St. Louis, St. Louis, MO 63110

**Author notes:** **Correspondence:** Warner BB,; Mitreva M. **Equal contribution:** Warner BB,; Rosa BA. **Email addresses** Barbara B Warner, Bruce A. Rosa, I. Malick Ndao, Phillip I. Tarr, J. Philip Miller, Sara K. England, Joan Luby, Cynthia Rogers, Carla Hall-Moore, Renay E. Bryant, Jacqueline D. Wang, Laura Linneman, Tara A. Smyser, Christopher D. Smyser, Deanna M. Barch, Gregory E. Miller, Edith Chen, John Martin, Makedonka Mitreva.

**Keywords:** infant, neonatal, parental, gut microbiome, social disadvantage, psychological stressors, machine learning, predictive modeling

## Abstract

Social disadvantage (SD) and psychological stressors (PS) trap some populations in poverty, resulting in health inequities. How these two factors become biologically embedded and the pathways leading to adverse health outcomes is unclear, especially in infants exposed to psychosocial adversity in utero and during early life. Variation in gut microbiome structure and functions, and systemic elevations in circulating cytokines levels as indices of inflammation, offer two possible causative pathways. Here, we interrogate the gut microbiome of mother-child dyads and maternal inflammatory markers, and compare high-SD/high-PS dyads to pairs with low-SD/Low-PS, and demonstrate that the GM of high-SD and high-PS mothers may already be compromised, resulting in the lowest observed inter-individual similarity in that group. The strong predictors of maternal high-SD and high-PS based on mothers and children microbiomes were phylogenetically very distinct bacteria indicating different GM pathways associated with SD versus PS. We identified sets of SD- and PS-discriminatory metabolic pathways in the mothers and in the children, however their predictive power was lower compared to the discriminatory bacterial species. Prediction accuracy was consistently greater for IL-6 than for the other inflammatory markers, supporting an association between systemic inflammation and psychosocial adversity. The gut microbiome of the infants can be used to predict the psychosocial adversity of mothers, and are embedded in the gut microbiota of 4-month-old infants.

## INTRODUCTION

Health inequities experienced by socially disadvantaged populations is an increasing and urgent societal issue [1, 2]. The psychosocial factors contributing to these disparities often begin early in life and include the prenatal environment which has profound effects on fetal and infant outcomes that can last a lifetime [3-5]. How social disadvatage (SD) and psychological stress (PS) becomes biologically embedded, and then leads to disparete health outcomes remains unclear.

The gut microbiome (GM) is one candidate driver of adverse outcomes. Many SD-related morbidities are associated with systemic chronic inflammation [6], and the GM, by shaping and modulating the immune system [7, 8], is associated with systemic inflammation and autoimmune disease, including many disorders associated with populations in which SD is widespread, and include diabetes [9], obesity [10], cardiovascular disease, and neurologic disorders [11].

The GM is itself shaped primarily by environment, with influences including environmental exposures of diet, antibiotics, exercise, stress and sleep deprivation [12]. Many of these exposures have distinct characteristics for individuals living with economic and psychosocial hardships. Despite increasing awareness of an intersection between the GM and psychosocial inequities [13, 14], few human studies [15-17] have examined the impact on gut microbial community structure and function, particularly in the perinatal period. Studies in pregnant women have focused on either maternal psychological state [18-22] or socioeconomic status [23-25] but not both, have used 16S RNA analysis, which limits, taxanomic and metabolic pathway identification, and have not linked mother-infants samples to examine maternal transfer within this context.

This study aims to fill these knowledge gaps. Using a prospective cohort assembled prenatally, we identify the distinct impact of exposure to SD and PS on GM structure and functions for mothers and their infants. We identify discriminatory taxa driving this association using whole metagenomic shotgun (WMS) sequencing, and attempt to relate systemic inflammation to the GM by examining maternal prenatal serum cytokines. Understanding the interaction of the gut microbiome with social determinants of health holds great promise for future preventions due to its potential alterability and plasticity over time. This is particularly true for the perinatal period, a critical developmental window in which perturbations in GM community structure and functions have long-lasting effects [26-28].

## RESULTS AND DISCUSSION

A subset of 121 mother–child dyads were drawn from the larger parent study, Early Life Adversity Biological Embedding and Risk for Developmental Precursors of Mental Disorders (eLABE), which enrolled mothers prenatally and followed their infants after brith. These 121 mothers were drawn from across the psychosocial spectrum and representative of the parent cohort with average ages of 29.8 (± 5.1 years) vs. 29.2 (± 5.3 years), similar dietary Healthy Eating index profiles 59 (± 10.6%) vs 58 (± 9.9%), and gestational age at delivery 39.0 (± 1.1wks) vs. 38.3 (± 2 weeks). Mothers and their children were classified based on composite scores from two latent constructs, SD and PS [29]. The SD construct included 7 individual domains across income, neighborhood deprivation, insurance, and education. The PS construct included 9 individual domains across depression, stress, short term and lifetime, and racial discrimination (*see* Methods for details). To define an effect of SD from that of PS on the maternal and children microbiome, we examined the interrelationships among pre- and post-natal adversity and circulating cytokines as biomarkers of inflammation, with the GM. The mothers’ GM was profiled using third trimester stool samples before parturition (average -18.0 ± 16.9). Their children’s stools at were sampled 130.4 (± 13.1) days after birth. Circulating cytokines were analyzed in maternal blood samples obtained during the 3^rd^ trimester of pregnancy. Participant characteristics are provided in **Table 1** and **Supplementary Table S1**.

**Table 1.**
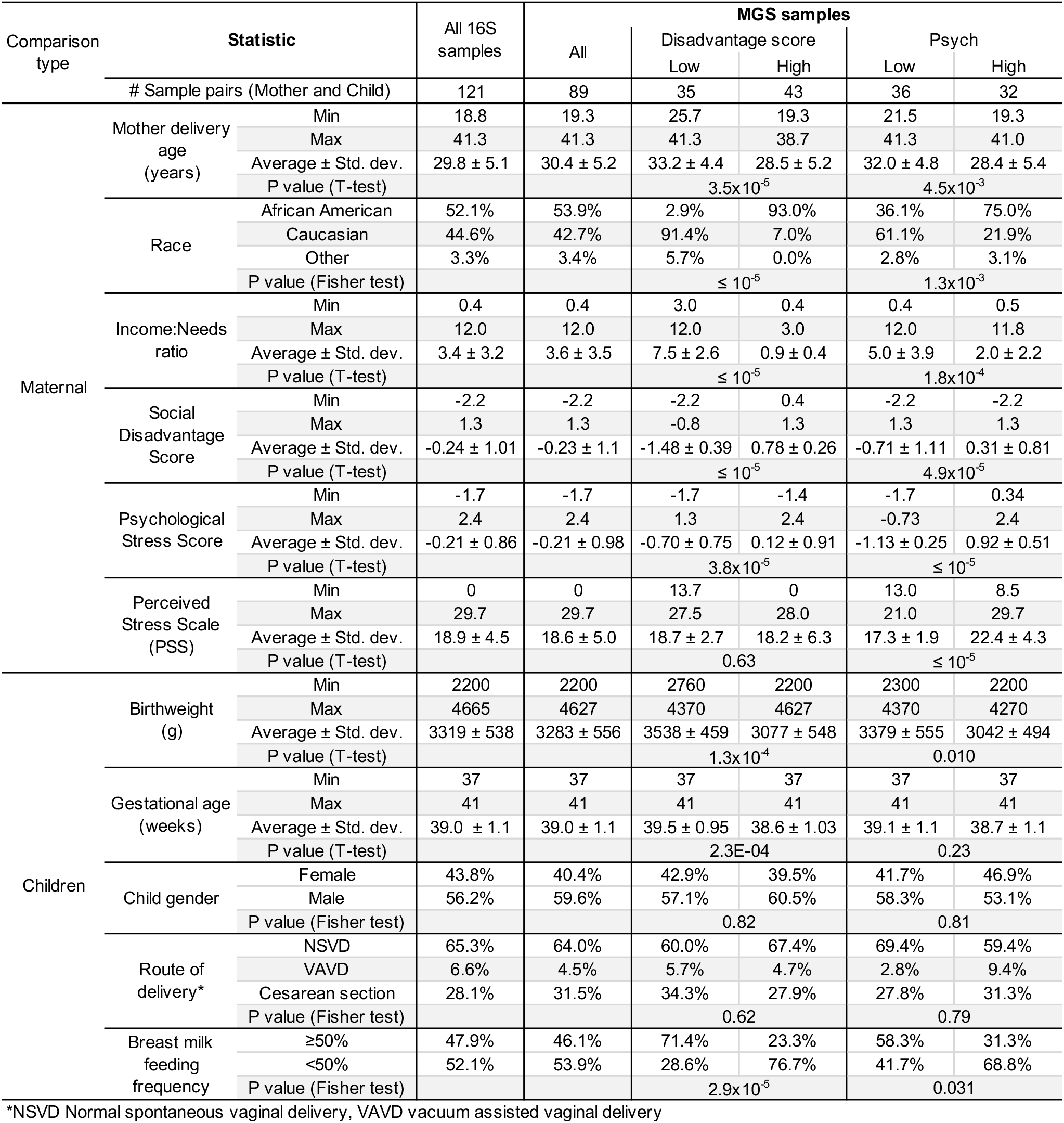
Patients characteristics at study entry for each of the primary comparisons of interest. “Low” and “High” Disadvantage and Psych scores are separated according to the distribution of the metadata, as shown in **Figure 1**. Complete characteristics of all samples organized per individual sample is available in **Supplementary Table S2**.

### Maternal SD and PS relation to maternal and child pre- and post-natal GM structure and function, based on high level taxonomic profiling

We performed targeted metagenomic sequencing (*see* Methods) for 121 mother-child dyads (**Figure 1**). Among the 121 mother–child dyads (**Figure 1A**), the SD scores were positively correlated with PS scores (r = 0.394, *P* < 10^−5^; **Figure 1B**), corroborating the literature [30] and demonstrating the importance of differentiating effects from these two overlapping variables. Several variables, including race, breast milk feeding frequency, healthy eating index and income-to-needs ratio, were significantly associated with high-vs-low SD and PS scores, also expected because these variables were used as inputs to calculate these scores [29] (**Table 1; Supplementary Table S1**).

**Figure 1:**
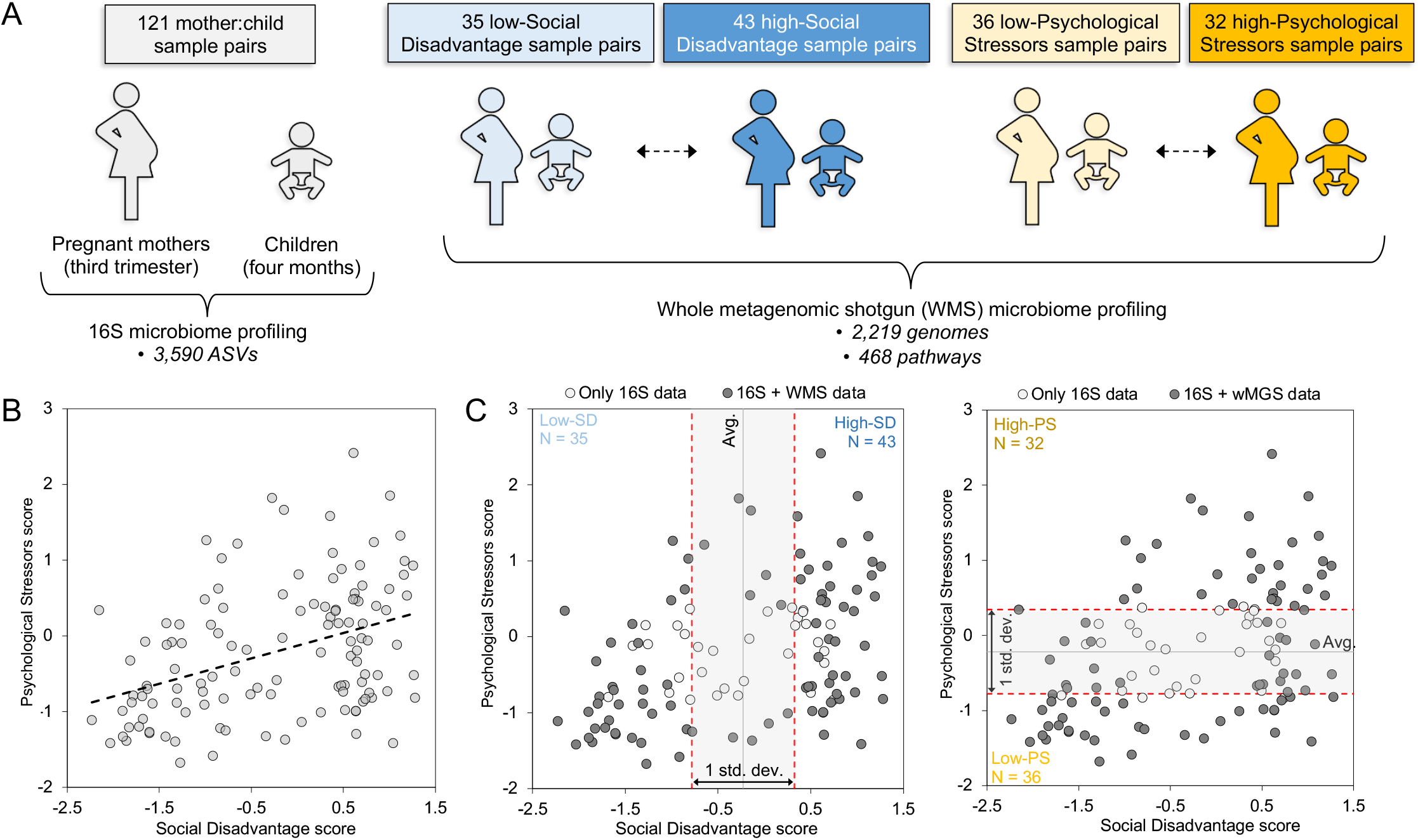
Overview of cohorts. (**A**) Portrayal of the 16S rRNA and WMS sample sets. (**B**) Social Disadvantage (SD) scores are positively correlated with Psychological Stressor (PS) scores (across the 121 sample pairs). (**C**) The 89 WMS samples are divided into “low-SD”, “high-SD”, “low-PS” and “high-PS” groups based on the distribution of values (samples within 1 standard deviation of the average value for each variable are excluded).

Across all samples, 3,072 amplicon sequence variants (ASVs) representing unique 16S nucleotide sequences were detected. Relative abundance values per ASV per sample are provided for all 242 samples in **Supplementary Table S2**, and nucleotide sequences for each identified ASV are provided in **Supplementary Table S3**. In the mothers, SD and PS scores had an overall negative correlation with α-(within sample) diversity based on the bacterial taxonomic composition, although this correlation was not significant (**Figure 2A**), consistent with previous reports of decreased α-diversity associated with lower Socioeconomic Status (SES) [15-17]. In contrast, among the children, α-diversity was positively correlated with SD (r = 0.581, *P* < 10^−5^) and PS scores (r = 0.350, *P* = 8.3×10^−5^) (**Figure 2B**). This observation may be partly explained by the lower frequency of breast feeding in the high-SD mothers (**Figure 2B)**, as it is reported that breast-fed infants had lower GM α-diversity compared to formula-fed infants at three months of age, but the GM diversity increased by six months [31].

**Figure 2:**
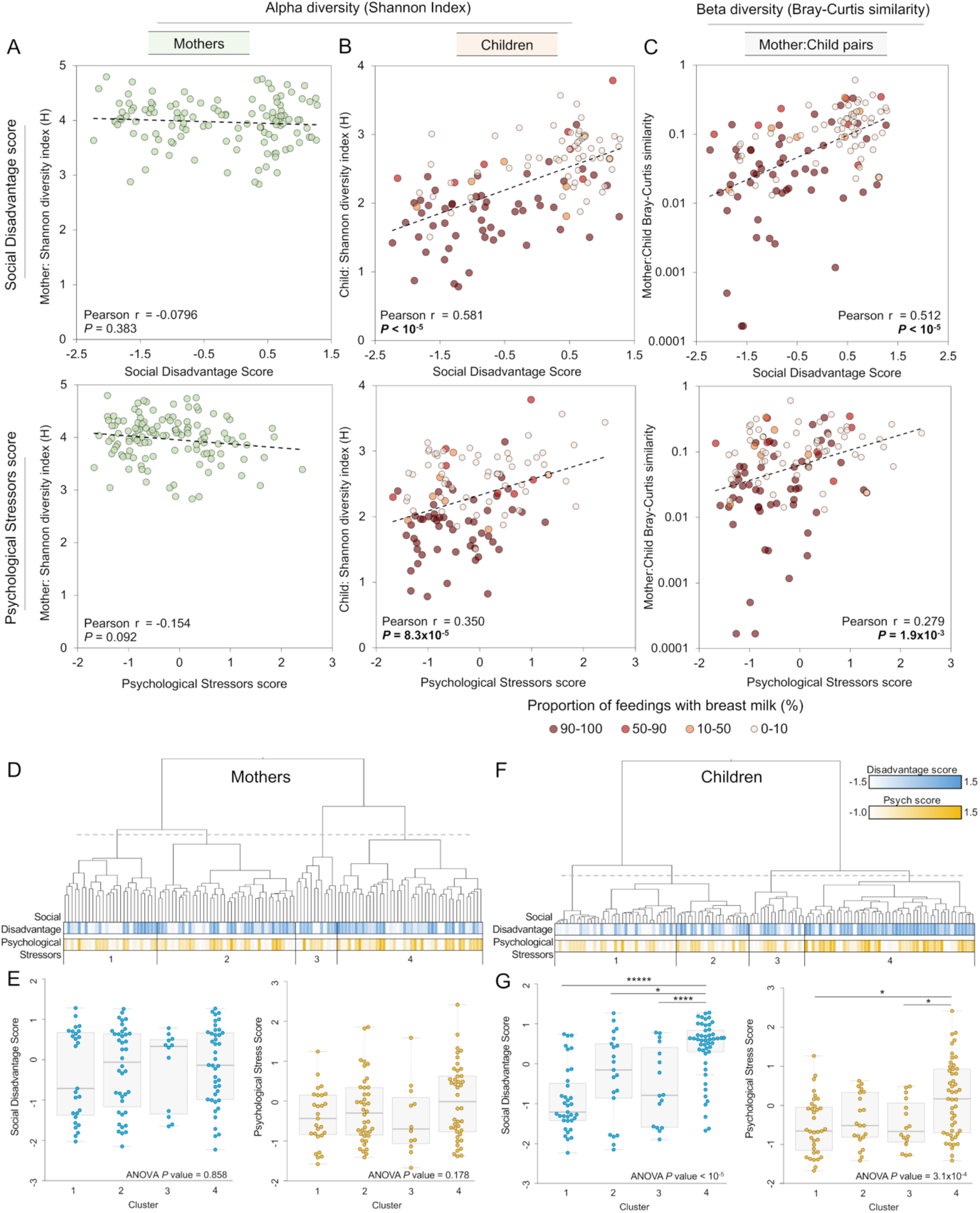
GM sample diversity and composition comparisons with Social Disadvantage (SD) and Psychological Stressors (PS) scores. (**A**) SD scores and PD scores do not significantly correlate with GM α-diversity (Shannon diversity index) in the 121 stool samples from mothers. (**B**) α-diversity in the children is positively correlated with SD (*P* < 10^−5^) and PS (*P* = 8.3×10^−5^) scores. Relative proportions of human milk feeding are also shown, to provide additional context for potential sources of differential diversity. (**C**) β-diversity, measured by Bray-Curtis dissimilarity between the GM of each mother-child dyad, is positively correlated with SD (*P* < 10^−5^) and PS (*P* = 1.9×10^−3^) scores. (**D**) Hierarchical clustering (based on Bray-Curtis dissimilarity across all ASVs, complete linkage) identifies four major GM profile-based clusters in the mother. (**E**) No significant differences in SD or PS scores were identified between the four clusters based on maternal GM profiles (one-way ANOVA). (**F**) Hierarchical clustering identifies four major GM clusters in the children. (**G**) Among the GM clusters in children, cluster 4 has greater SD (one-way ANOVA *P* < 3.4×10^−5^) and PS (3.1×10^−4^) scores. Tukey post-hoc tests were used to identify significant differences between each cluster, \**P* ≤ 0.05, **** P ≤ 10^−4^, *****, P ≤ 10^−5^.

An examination of the similarity between mother and child GM (β-diversity) measured by the Bray-Curtis statistics, showed that mother–child dyads with low-SD scores had significantly less similarity (*P* < 10^−5^, **Figure 2C**). This may be partially due differences in α-diversity, where formula-fed/high-SD infants have GM that are more similar to their mother’s GM because of greater overall α-diversity. Bray-Curtis dissimilarity-based clustering identified significant associations between SD scores and PS scores in the microbiome profiles in the children, but not in the mothers (**Figures 2D-2G**). We identified four major GM clusters in the mothers, but none significantly differ in SD or PS (**Figures 2D** and **2E**). We also identified four major sample clusters in children, with cluster 4 having significantly greater SD scores than the other three clusters (ANOVA *P* < 10^−5^), and also higher PS scores than clusters 1 and 3 (ANOVA *P* = 3.1×10^−4^) (**Figures 2F** and **2G**). These results suggest that high-SD and high-PS scores are significantly associated with a distinct overall GM profile in the children, but not in the mothers.

We next compared Bray-Curtis similarity between each sample dyad, within and between high and low SD and PS groups (**Figure 3**). High-SD mothers had significantly more variable microbiome profiles than those of low-SD mothers, who presumably had more consistent healthy microbiome profiles. This effect was even stronger for the PS comparison, with low-PS mothers having the most similar GM to each other than in any of the other comparisons, and the high-PS mothers having as similar between-sample diversity as the low-SD mothers (**Figure 3A**). High-SD children had the most similar microbiome profiles (**Figure 3B**), which may relate to their increased α-diversity (**Figure 2B**). Low-SD children also have some overall similarity in their GM profiles, but the low-SD and high-SD children GMs have little similarity (consistent with clustering results in **Figure 2F**). The same is true for PS in the children, except the within-group similarity for low-PS and high-PS were not significantly different. Overall, these results suggest that high-SD and high-PS mothers have divergent, variable microbiomes compared to low-SD and low-PS mothers, who share some commonality in overall GM profiles. Taken together, these results suggest that high-SD and high-PS mother already have compromised GM resulting in low observed inter-individual similarity compared to the low-SD and low-PS group, which has a more similar and healthier mature GM.

**Figure 3:**
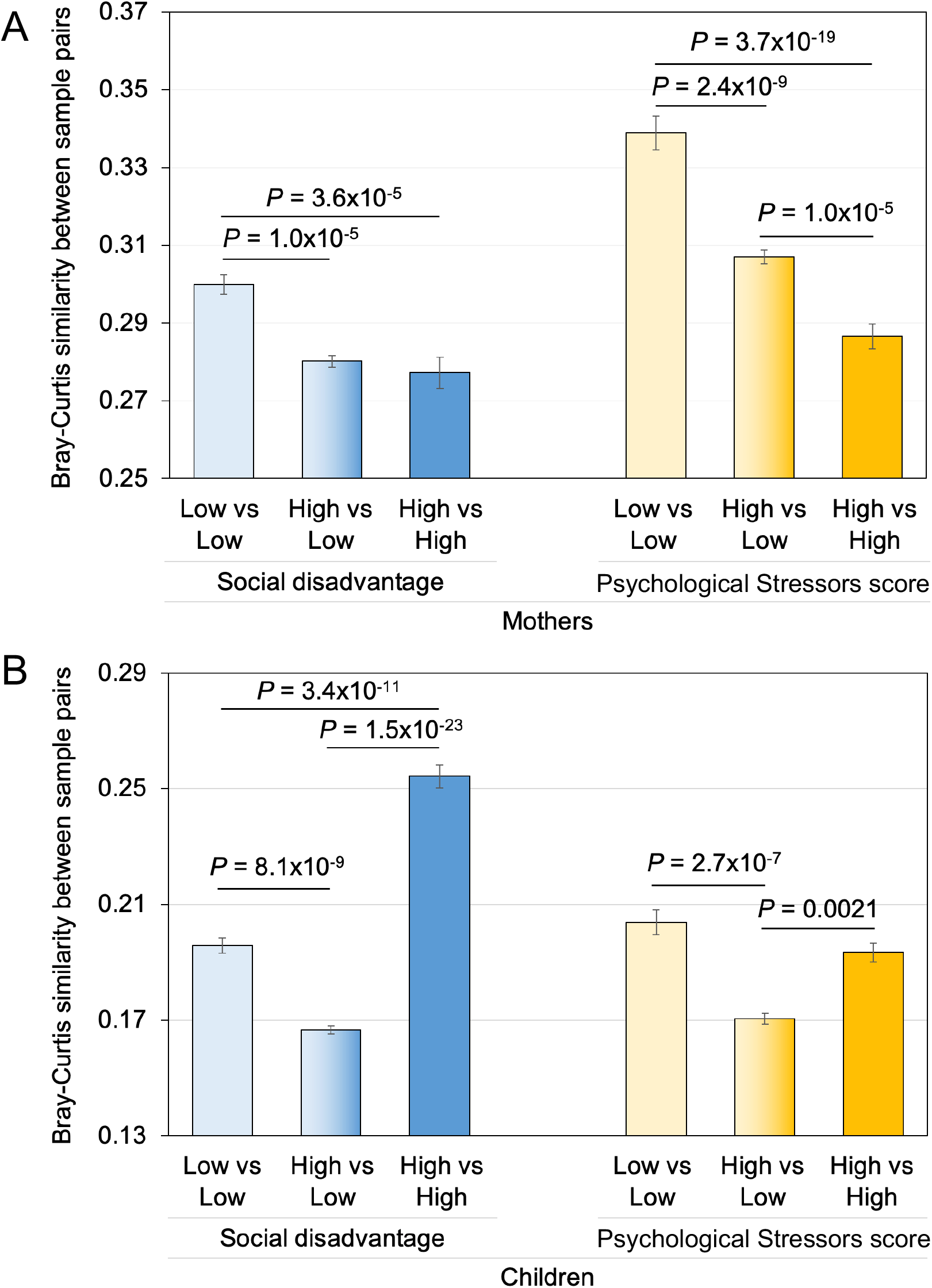
Comparisons of β-diversity between sample sets based on high-vs-low SD and PS, quantified by Bray-Curtis similarity between sample pairs, and FDR-corrected two-tailed T-tests with unequal variance used to test significance (**A**) Comparisons of within- and between-group β-diversity of the GM for mothers with high-SD, low-SD, high-PS and low-PS. (**B**) Comparisons of within- and between-group GM β-diversity in children with high-SD, low-SD, high-PS and low-PS.

### Species-level gut microbiome profiles and metabolic pathway reconstructions of 89 mother–child dyads

We performed WMS sequencing on a subset of stools from 89 of the 121 mother–child dyads, selected based on the distribution of SD and PS scores (**Figure 1C**) (greater and less than the average value + 0.5 and – 0.5 standard deviations). The profiles of the 178 GMs (89 mothers and their 89 children), were generated from an average of 6Gb reads per sample, characterized using the Unified Human Gastrointestinal Genome (UHGG) [32]. Relative abundance values per species per sample were calculated using normalized depths [33] for downstream analysis. A total of 2,219 bacterial genomes were detected across all samples (**Figure 4A**). Genome taxonomic annotations and relative genome abundance values per sample are provided for all 178 samples (**Supplementary Table S2)**. Twelve genomes were detected frequently across all mothers and all children samples (≥ 40% of both), including five *Bifidobacterium* genus members (*B. infantis, B. breve, B. bifidum, B. catenulatum* and *B. adolescentis*), with *Bifidobacterium infantis* being the most frequently detected genome across all samples (69.7% of mothers and 89.9% of children). These 12 also included two *Bacteroides* species (*B. dorei* and *B. xylanisolvens*), two *Faecalicatena* species (*F. gnavus* and *F. unclassified*), *Flavonifractor plautii* and *Eggerthella lenta* (**Figure 4B**). Four of these species were also identified as “core mother-infant shared species” in a previous WGS study [34], however the limited overlap may be in part a result of using different genome reference databases (identifying clade-specific marker genes from MetaPhlAn2 [34, 35] vs mapping to the Unified Human Gastrointestinal Genome (UHGG) [32]) in these 2 studies.

**Figure 4:**
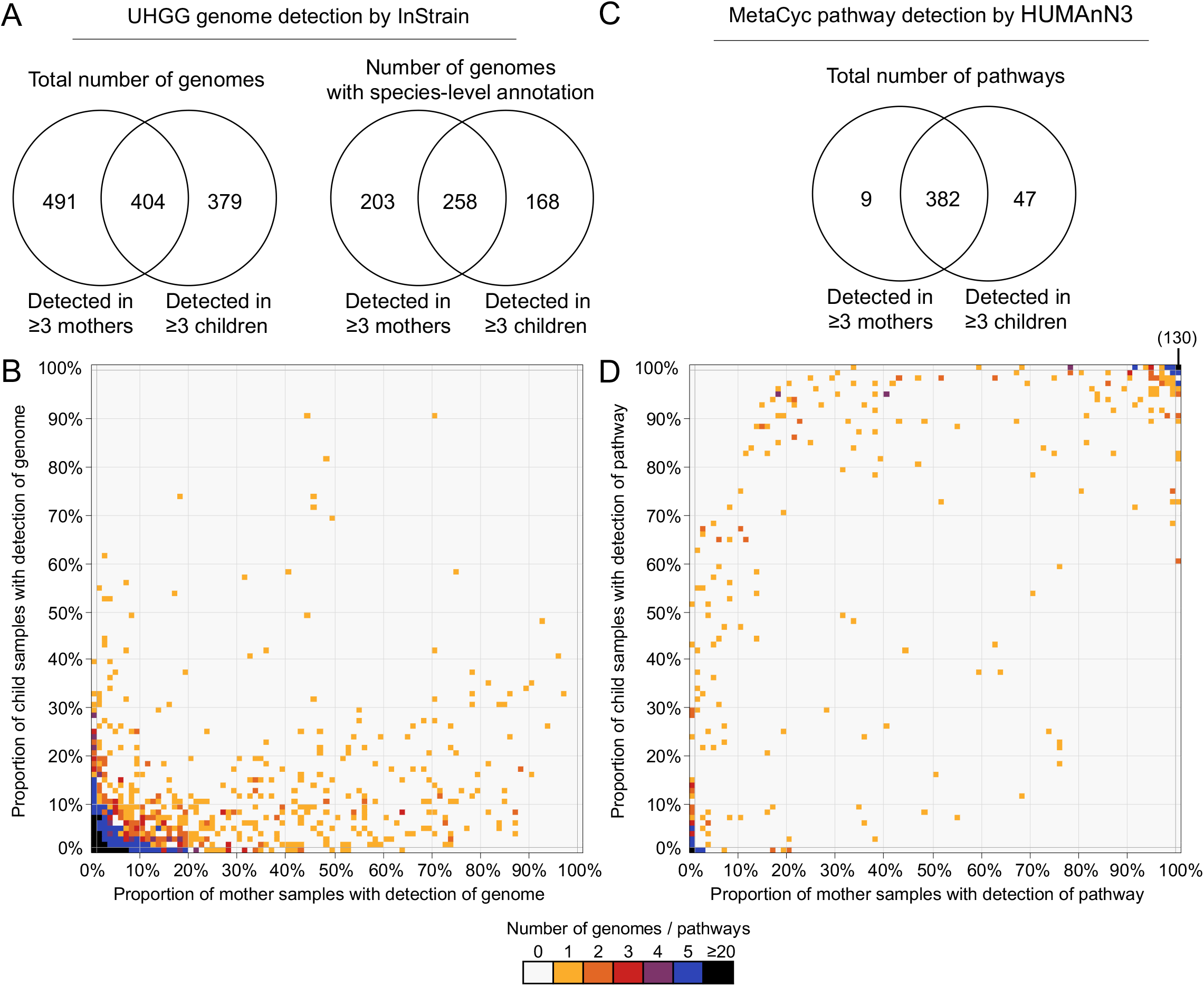
Genome and pathway detection in the 89 mother and 89 child WMS samples. (**A**) The total number of bacterial genomes and genomes with species-level taxonomic annotation detected in at least 3 maternal samples and/or at least 3 child samples. (**B**) For each of the 2,219 bacterial genomes detected in any sample, the proportion of mother and child samples that were detected in the dataset. (**C**) The total number of metabolic pathways detected in the GM from at least 3 mother and at least 3 child stools. (**D**) For each the 468 metabolic pathways detected in any sample, the proportion of mother and child samples with detection in the dataset.

We next reconstructed the metabolic pathways to compare the functional potential of the GM communities (based on the relative abundance of metabolic pathways [36] in read counts per million, CPM) using HUMAnN3 [37-39]. A total of 468 pathways were detected across all samples, 438 of which were detected in at least 3 mother or 3 children GMs (**Figure 4C**). Relevant pathway annotations and relative pathway abundance values per sample are provided for all 178 samples (**Supplementary Table S2)**. In contrast to the genomes with relatively sparse identification across samples, almost half (46.3%) of all detected pathways were identified in ≥ 90% of samples in both the mothers and the children (top right of the plot, **Figure 4D**), including 130 pathways (27.8%) detected in all 178 samples. Despite the taxonomic differences between the GM of mothers and children (31.71% shared genomes; **Figure 4A**), the core set of microbial functions is conserved across the groups with 87.2% of the pathways being encoded by both mothers and children’s GMs (**Figure 4C**). This greater similarity between mothers and children in their microbial metabolic pathways compared to their microbial taxonomic profiles has been observed previously, and may be attributed to “core” microbial community functions essential for all species, despite the distinct populations of species adapted to different diets at different stages of life [34].

### Bacterial species discriminating between mothers SD and PS

To dissociate the impact of the highly inter-related SD from PS on the maternal GM, and to identify discriminatory bacterial taxa that are strong predictors of mother’s SD and PS scores, we analyzed taxonomic and pathway GM profiles using two statistical approaches. First, we used supervised Random Forest (RF; [40]) machine-learning was used with a two-round approach [41] to (i) quantify the ability to predict SD and PS classification based on the mothers’ and the childrens’ microbiome profiles (**Figure 5**), and (ii) identify the specific genomes and pathways that most strongly differentiate between the high and low SD and PS scores (**Figures 6** and **7**; *see* Methods). Second, we used linear discriminant analysis effect size (LEfSe; [42]) to test differential genome and pathway abundance testing, to assign *P* values and “effect size” values for significance, as additional validation of RF results. Differential abundance statistics for all genomes and all comparisons are provided in **Supplementary Table S4**.

**Figure 5:**
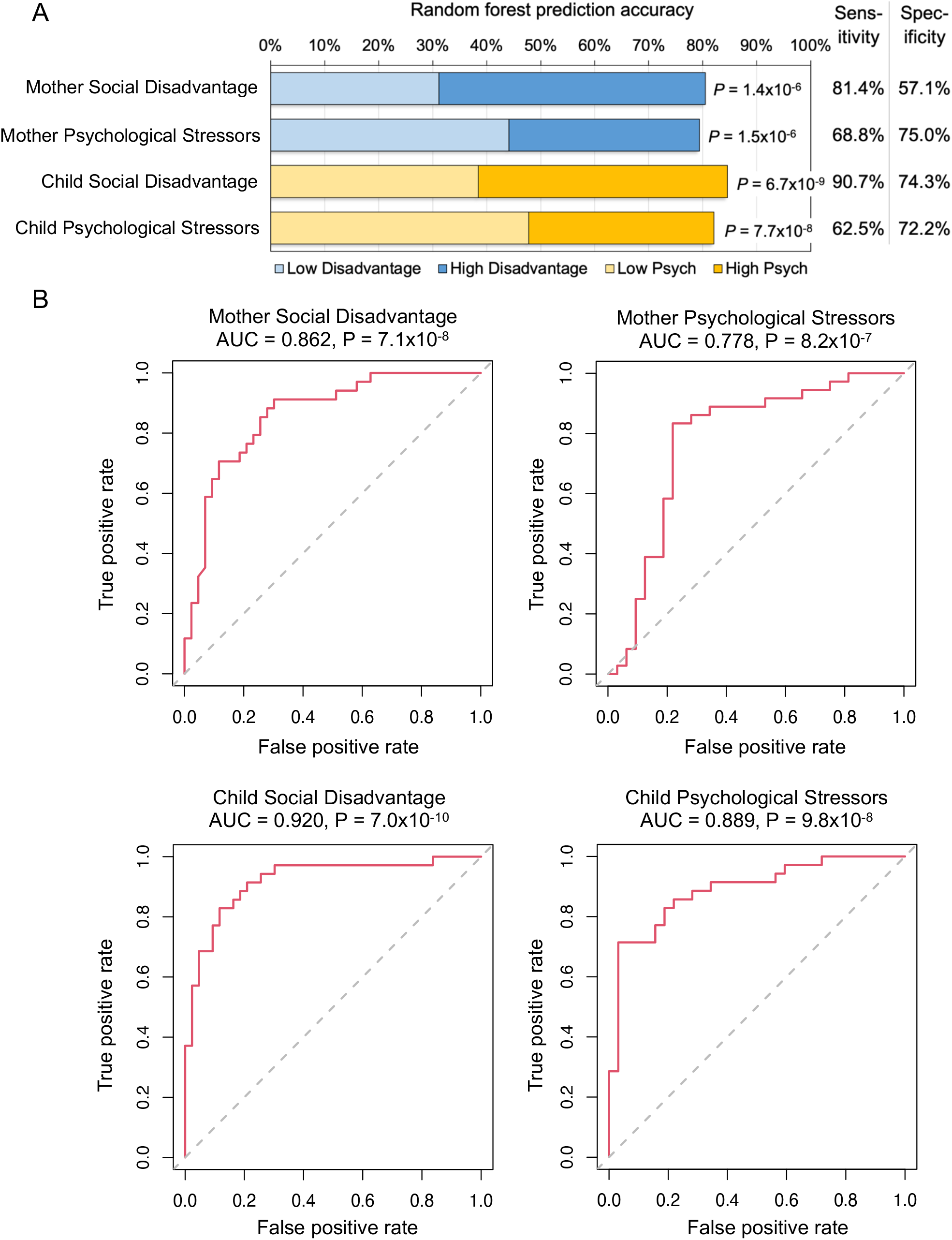
The Random Forest (RF) classification accuracy (low-SD vs high-SD and low-PS vs high-PS, out-of-bag error) based on relative UHGG genome abundance in the mothers and children. (**A**) Overall classification accuracy, with *P* values indicating significance based on FDR-corrected binomial distribution tests (compared to random sample assignment). (**B**) For each of the four classification tests, receiver operating characteristic (ROC) curves are shown based on RF models, with the area under the curve (AUC) scores and associated Wilcoxon rank sum test results for each ROC curve indicated.

**Figure 6:**
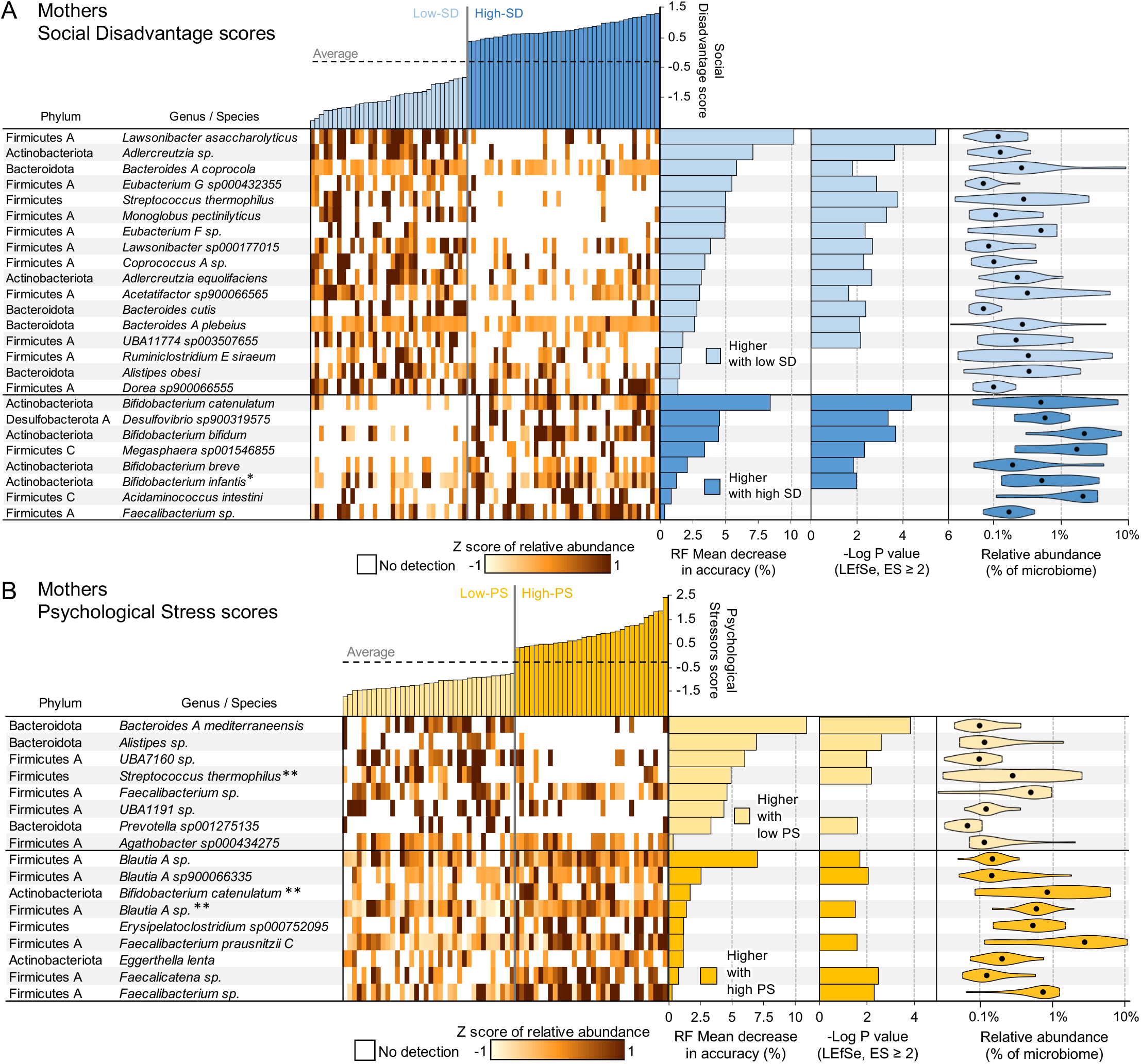
WMS genome differential abundance in the mothers, based on comparisons of high-vs-low SD scores and high-vs-low Psych scores. (**A**) Based on WMS genome profiles, RF can successfully classify mothers as having low and high SD 80.5% of the time (*P* = 1.4×10^−6^ compared to random assignment; FDR-corrected binomial distribution test). Taxonomy and relative abundance per sample for the genomes with the highest predictive value in the top-25 RF model are shown (ranked by mean decrease in accuracy of the RF model; MDA). Also displayed are -Log of the Kruskal-Wallis test P values from LEfSe (no value shown if the effect size was <2) and the overall abundance of the taxa when present. (**B**) Based on WMS genome profiles, Random Forest (RF) can successfully classify mothers into high and low Psych groups 79.4% of the time (*P* = 1.5×10^−6^). *Associated with low-SD in the children, **associated with high Psych in the children.

We identified a set of SD and PS-discriminatory bacteria (**Figure 6)** whose relative abundances can classify mothers into low- or high-SD 80.5% of the time (*P* = 1.4×10^−6^ compared to random assignment; FDR-corrected binomial distribution test; AUC = 0.862, *P* = 7.1×10^−8^, Wilcoxon rank sum test), and as low- or high-PS 79.4% of the time (*P* =1.5×10^−6^; AUC = 0.778, *P* = 8.2×10^−7^; **Figure 5**). The best predictors of SD (**Figure 6A**) and PS (**Figure 6B**) scores in the mothers were identified according to mean decrease of accuracy (MDA) scores (*see* Methods) for each genome across the low-SD group and the high-SD groups. The microbiome signature of low-SD and low-PS samples consisted of bacterial species present in high abundance in most of the samples, but which were detected with zero or very low abundance in the high-SD or high-PS samples (**Figure 6**).

Low-SD mothers are identified by increased abundance of many Firmicutes A genomes (**Figure 6A**). *Lawsonibacter asaccharolyticus*, a recently-identified butyrate-producing species [43, 44], was the strongest predictor of low-SD scores in the mothers (MDA = 10.2%, LEfSe effect size = 2.7, *P* = 3.7×10^−6^). GM-derived butyrate has a wide range of beneficial effects on health including regulating fluid transport, reducing inflammation, reinforcing the epithelial defense barrier, and modulating intestinal motility via mechanisms that include potent regulation of gene expression [45], but *L. asaccharolyticus* has not been previously and independently associated with these beneficial effects. *Streptococcus thermophilus* was also among the top 5 predictors (MDA = 5.0%, LEfSe effect size = 3.2, *P* = 1.7×10^−4^) and is used to produce yogurts and cheeses, has properties beneficial to health including the prevention of chronic gastritis and diarrhea, and immunomodulatory properties with possible benefits in inflammation [46]. *S. thermophilus* was also among the predictors of low-PS, and this species was detected with zero or low abundance in high-PS and high-SD individuals.

The predictors of high-SD and high-PS were phylogenetically quite distinct, enriched for Actinobacteria and Firmicutes C, vs. mainly Firmicutes A, respectively. Genomes from four *Bifidobacterium* species (*B. catenulatum, B. bifidum, B. breve* and *B. infantis*) are among the six most strongly associated with high-SD, and *B. catenulatum* is associated with high-PS as well. It is recognized that *Bifidobacteria* in the human gut vary with age, and while quantitively some are particularly important in infant GM its presence with aging is stable but abundance changes over time. In general high abundance of *Bifidobacteria* is related to gut homeostasis and health maintenance and protection, in part by producing a number of potentially health promoting metabolites including short chain fatty acids, conjugated linoleic acid and bacteriocins, and *Bifidobacteria* is postulated to improve health [47]. However, qualitative and quantitative (increasing abundance) in *Bifidobacteria* are associated with inflammatory disorders such as diverticulitis, inflammatory bowel disease, and colorectal cancer [48]. Additionally, a recent review of GM variations associated with major depressive disorder (MDD) identified *Bifidobacterium* as one of three genera most consistently associated with MDD across studies [49]. While the specific functional role of these high-SD associated *Bifidobacteria* species is unclear, there is a striking increase in overall Bifidobacterial abundance in the high-SD mothers (**Supplementary Figure S1**).

*Bacteroides A mediterraneensis* was most strongly associated with low-PS in mothers (MDA = 10.9%, effect size = 2.5, *P* = 1.5×10^−4^; **Figure 6B**). In mice, stress exposure reduces abundance of *Bacteroides* in the GM [50], and in humans, *Bacteroides* is one of five genera associated with healthy status vs. MDD patients [51], the but this is the first report of *Bacteroides A mediterraneensis* specifically being associated with PS in a human cohort. The same human MDD study [51] also identified *Faecalibacterium* and *Prevotella* as being negatively associated with MDD, and in this study *Faecalibacterium sp*. and *Prevotella sp001275135* were the 5^th^ and 7^th^ strongest predictors to low-PS (respectively).

Three species of *Blautia sp*. were among the top four most strong predictors of high-PS in the mothers, and none of these four species were associated with SD, suggesting a specific link with psychological stressors. In human studies, *Blautia* was one of ten genera associated with MDD [51], and *Blautia* and *Eggerthella* (represented in the high-PS mothers by *E. lenta*) were significantly correlated with PSS scores [52]. The latter study also identified *Blautia* and *Bifidobacteria* (represented in the high-PS mothers by *B. catenulatum*) as being significantly associated with MDD [52]. However, we wish to note that the overall abundance of *Blautia* in the microbiome remained fairly consistent across the mothers (**Supplementary Figure S2**), and only the species identified in **Figure 6B** predict PS.

### Bacterial species in the infant GM discriminating prenatal maternal SD and PS

A set of SD and PS-discriminatory bacterial genomes correctly classify children into low- or high-SD in 84.6% of comparisons (*P* = 6.7×10^−9^ compared to random assignment; FDR-corrected binomial distribution test; AUC = 0.920, *P* = 7.0×10^−10,^, Wilcoxon rank sum test), and as low- or high-PS in 82.1% of comparisons (*P* =7.7×10^−8^; AUC = 0.889, *P* = 9.8×10^−8^) **Figure 5**). The top predictors of SD score (**Figure 7A**) and PS score (**Figure 7B**) in the children were identified according to MDA scores.

**Figure 7:**
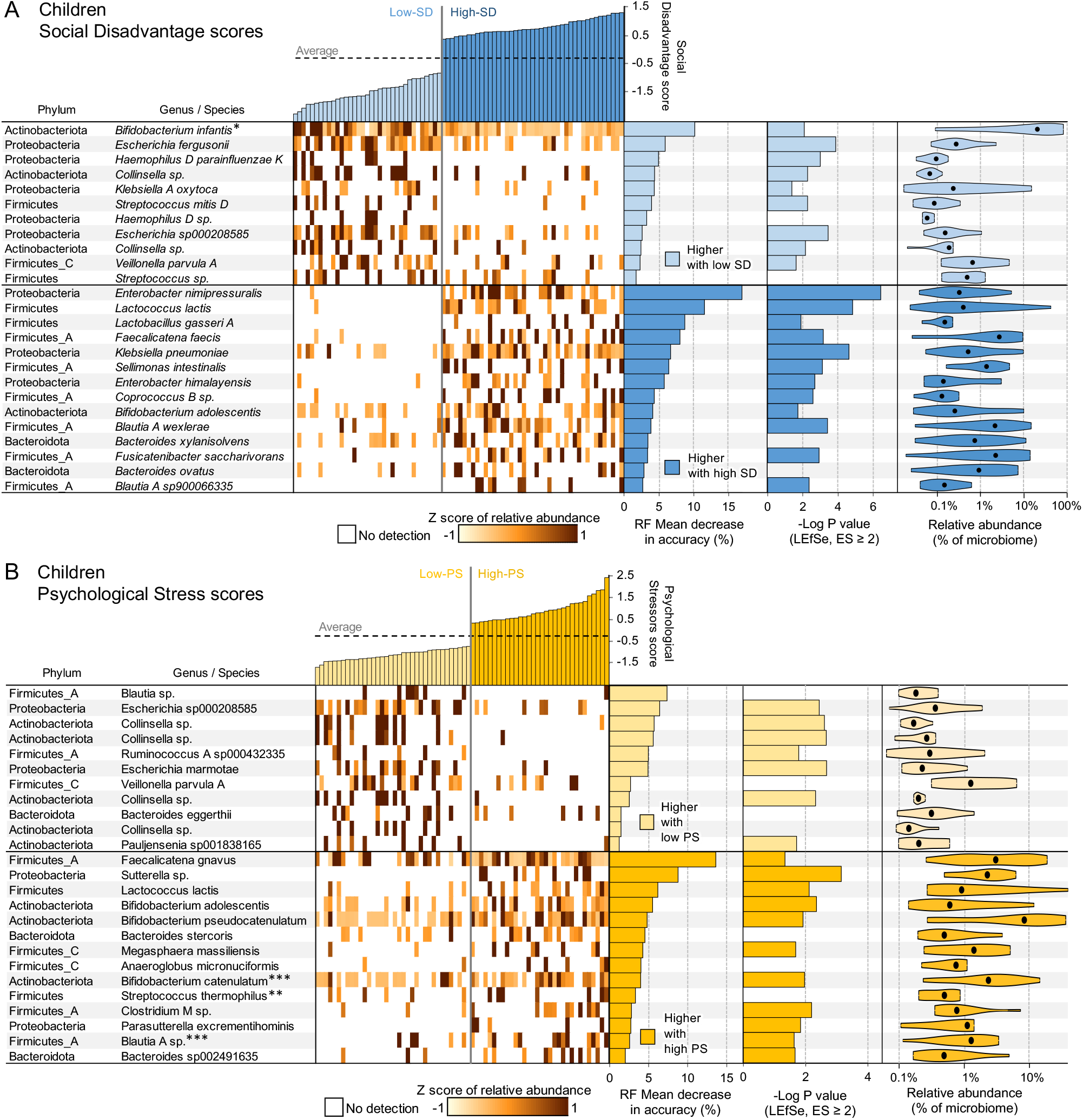
WMS genome differential abundance in the children, based on comparisons of high-vs-low Social Disadvantage (SD) scores and high-vs-low Psychological Stressors (PS) scores. (**A**) Based on WMS genome profiles, Random Forest (RF) can successfully classify children as low- and high-SD 84.6% of the time (*P* = 6.7×10^−9^ compared to random assignment; FDR-corrected binomial distribution test). Taxonomy and relative abundance per sample for the genomes with the highest predictive value in the top-25 RF model are shown (ranked by mean decrease in accuracy of the RF model; MDA). Also displayed are -Log of the Kruskal-Wallis test P values from LEfSe (no value shown if the effect size was <2) and the overall abundance of the taxa when present. (**B**) Based on WMS genome profiles, RF can successfully classify children into high and low-PS groups 82.1% of the time (*P* = 7.7×10^−8^). *Associated with high-SD in the mothers, **associated with low Psych in the mothers, ***also associated with high-PS in the mothers.

Among the predictors that are most important for classification to high-SD in children and with the largest effect size are *Enterobacter nimipressuralis*, nearly absent in low-SD children (detected in only one sample), and *Klebsiella pneumoniae*, both proinflammatory lipopolysaccharide expressing Proteobacteria [53]. The strongest predictor of children with low-SD was *B. infantis*, a species frequently used as a probiotic to diminish inflammation and associated with breastfeeding [54]. *B. infantis* represented an average of 28.3% of the GM of the low-SD infants, but only 5.2% of the GM in the high-SD infants (MDA = 10.14%, LEfSe effect size = 5.1, *P* = 0.0083; **Supplementary Figure S1**). The top taxa based on the comparison of the SD scores in the mothers were not proinflammatory and belonged to genomes from four *Bifidobacterium* species previously associated with several putatively beneficial metabolites [47].

There was an overlap of the predictors for low-SD and low-PS, including *Veillonella parvula A*, and several *Collinsella* spp. *Veillonella* is a signature taxa of the 4-month microbiome, and with *Collinsella* have been found in breast-fed microbiome indicating reduced concentration of oxygen, increased production and utilization of lactic acid which is specific for milk dominated diet [55]. The high-SD and high-PS-discriminating bacteria were enriched for a broad range of evolutionarily distinct Firmicutes species (**Figure 7**). The best high-PS predictors included *F. gnavus* reported as pathobiont associated with inflammatory bowel disease [56] and *Sutterella* sp. *Sutterella* species are prevalent commensals in the human GM with mild-proinflammatory properties [57].

### Metabolic pathways associated with SD and PS in mothers and children

The relative genomic abundance data from the WMS dataset has more features and more taxonomic specificity than metabolic pathway abundance data, because pathways are much more conserved across samples (**Figure 4**). Genomic data therefore enables better estimation of overall GM associations with SD and PS scores (**Figure 5**). Nevertheless, the same statistical approaches (*see* Methods; RF and LEfSe) were used to identify sets of SD- and PS-discriminatory metabolic pathways in the mothers and in the children (defined here using MetaCyc, a curated database of experimentally elucidated metabolic pathways from all domains of life [36]). Differential abundance statistics for all pathways and all comparisons are provided in **Supplementary Table S5**.

Based on metabolic pathway profiles, machine learning correctly classifies mothers as high- and low-SD in 71.8% of comparisons (*P* = 8.5×10^−4^) and as high- and low-PS in 69.1% of comparisons (*P* = 0.0023). The top predictive pathways for both comparisons are shown in **Table 2**. The pathways with greatest association with high-SD in mothers include three related to carbohydrate degradation (sucrose degradation IV, glycogen degradation I, starch degradation III), which might relate to the accompanying abundance of *Bifidobacterium* species, which are rich in carbohydrate metabolism pathways [58]. The “myo-, chiro- and scyllo-inositol degradation” pathway (PWY-7237) was most strongly associated with high-PS in mothers (MDA = 5.95, P = 2.1×10^−3^). Myo-inositol and chiro-inositol degradation by the gut microbiome contributes to inositol deficiency [59], which includes metabolic disorders involved with insulin function [59] and MDD when myo-inositol is deficient in the prefrontal cortex [60].

**Table 2.**
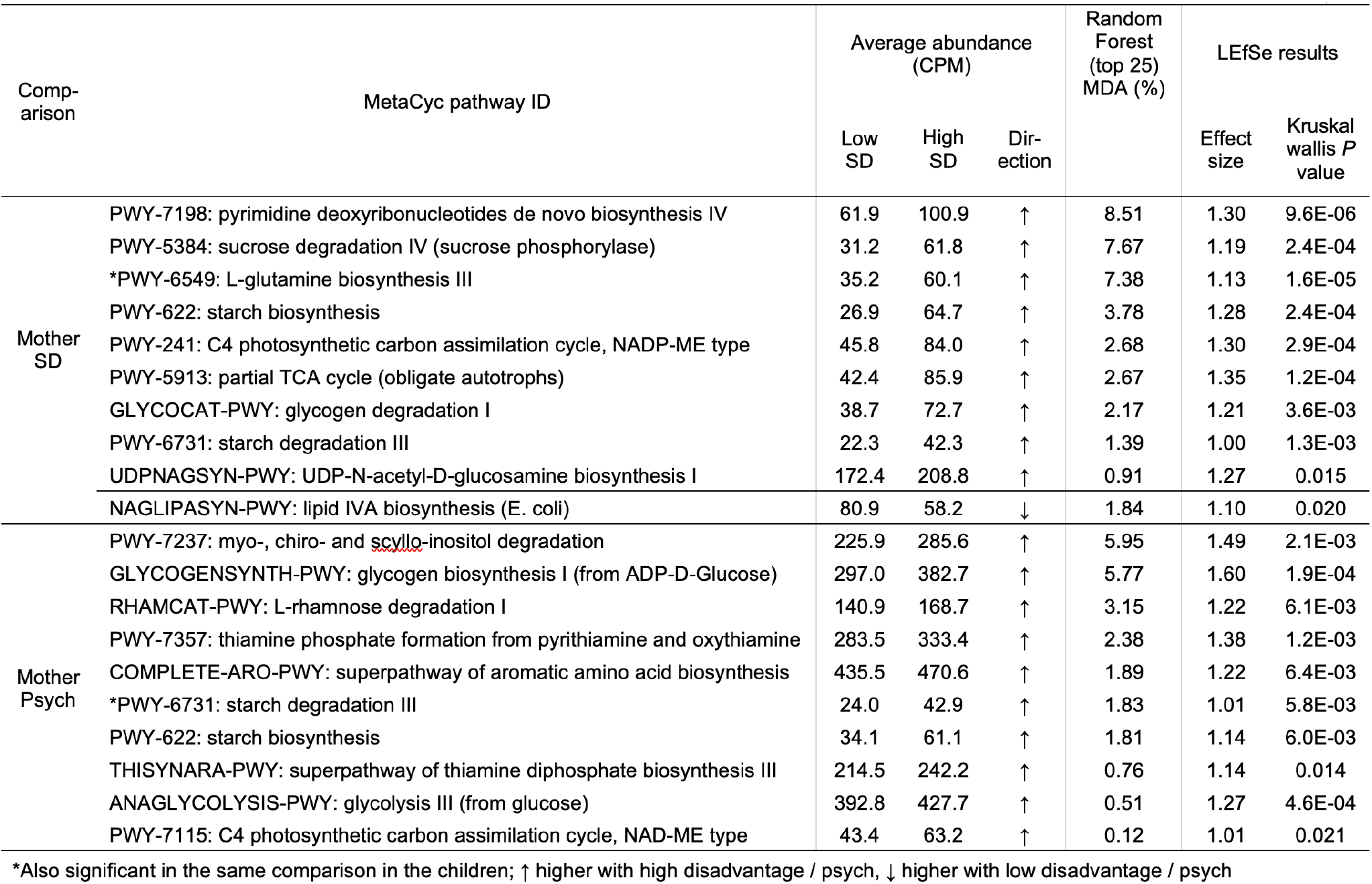
Metabolic pathways differential abundance in the mothers, based on comparisons of high-vs-low SD scores and high-vs-low PS scores. Machine learning algorithm can classify mothers as low and high SD 71.8% of the time (*P* = 8.6×10^−4^ compared to random assignment; FDR-corrected binomial distribution test). Average abundance and association (high or low) are shown for the pathways predictive value in the top-25 RF model (ranked by mean decrease in accuracy of the RF model; MDA), and with LEfSe effect size ≥ 1. The -log of the Kruskal-Wallis test *P* values from LEfSe is also shown.

Children are classified correctly as high- and low-SD in 85.9% of comparisons (*P* = 1.3×10^−9^) and as high- and low-PS in 72.1% of comparisons (*P* = 4.1×10^−4^) based on pathway abundance profiles. The top predictive pathways for both of these comparisons are shown in **Table 3**. Synthesis of L-glutamate and L-glutamine (PWY-5505) was the pathway most strongly associated with high-SD in the children (MDA = 7.9%, P = 7.9×10^−7^), and has been previously associated with obesity and visceral fat accumulation [61]. Here, L-glutamine biosynthesis was also associated with high-SD in both mothers and children. In supplementation studies of the gut microbiome, glutamine reduces the ratio of Firmicutes to Bacteroidetes and bacterial overgrowth or bacterial translocation, and increases the density of secretory immunoglobulin A (IgA) and IgA+ cells in the intestinal lumen [62].

**Table 3.**
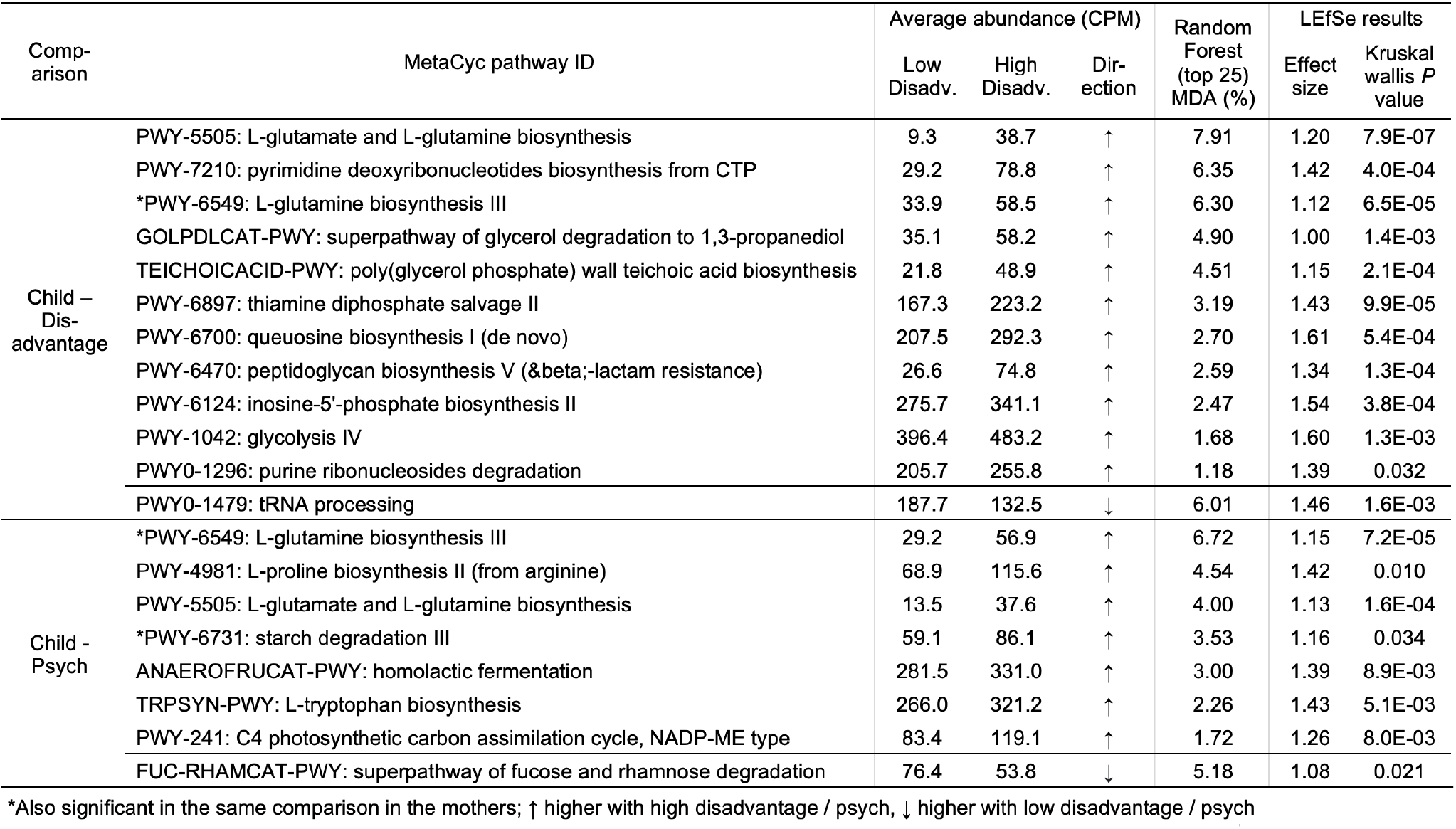
Metabolic pathways differential abundance in the children, based on comparisons of high-vs-low SD scores and high-vs-low Machine learning algorithm can correctly classify children as low and high mother SD 71.8% of the time (*P* = 8.6×10^−4^ compared to random assignment; FDR-corrected binomial distribution test). Average abundance and association (high or low) are shown for the pathways predictive value in the top-25 RF model (ranked by mean decrease in accuracy of the RF model; MDA), and with LEfSe effect size ≥ 1. The -log of the Kruskal-Wallis test *P* values from LEfSe is also shown.

### Bacterial species discrimination between SD and PS groups based on maternal circulating cytokines

The relationship between inflammation and the GM is examined through maternal systemic cytokines IL-6, IL-8, IL-10 and TNF alpha, measured across trimesters. In mothers, prediction accuracy of the GM was greatest for third trimester cytokine samples, consistent with timing of maternal stool samples. Prediction accuracy was consistently greater for IL-6 than for the other inflammatory markers (**Table 4**). Discriminatory bacterial genomes demonstrate the most consistent and greatest predictive accuracy for IL-6, which in preclinical models is centrally important in altering fetal brain development in maternal immune activation models, where placental inflammatory signals are relayed to the fetal brain [63-65]. We identified a set of discriminatory taxa from maternal third trimester stools, whose relative abundance successfully classify mothers into low- or high-IL-6 75.6% of comparisons (*P* = 2.2×10^−4^; **Supplementary Figure S3**) vs. random assignment (FDR-corrected binomial distribution test), and in 72.1% of comparisons of metabolic pathways (*P* = 2.2×10^−4^). In this case, high prenatal IL-6 concentrations are associated with the lower abundance of anti-inflammatory *Bacteroides* species, as evidenced by the RF and LEfSe significance (effect size 2.3, *P* = 0.017 for *Bacteroides A* and effect size 2.9, *P* = 0.02 for *Bacteroides faecis*), as reported in other inflammatory states [47, 66]. These data illustrate the importance of community balance, including presence and absence, on target outcomes, and the importance in the ability to identify these at species/strain levels. Interestingly, we also identified discriminatory taxa in the infant 4-month GM profile (**Figure 8**). Of all inflammatory marker comparisons, microbial genomes in children had the best overall accuracy for maternal third trimester IL-6 concentrations (high/low) based on relative abundance (77.8% accuracy, *P* = 6.0×10^−5^) and metabolic pathways (75.6%, *P* = 6.9×10^−4^). However, maternal IL-6 concentration was not significantly correlated with SD or PS in this subsample (**Supplementary Figure S4**), although higher SD is associated with higher maternal IL-6 in the full sample set. Thus, the relationship of the maternal GM to IL-6 is driven by taxa distinct from those identified in composite SD or PS values, but still have proinflammatory characteristics.

**Table 4.** Random forest (RF) machine learning predictive accuracy for high-vs-low inflammatory markers based on GM taxonomic and pathway profiles. Prediction accuracy was consistently greater for IL-6 than for the other inflammatory markers. Bolded values correspond to comparisons for which *P* < 0.005 (after FDR correction).

|  |  | Prediction accuracy |  |  |  | FDR-adjusted P value for prediction accuracy |  |  |  |
| --- | --- | --- | --- | --- | --- | --- | --- | --- | --- |
| | | IL-6 | IL-8 | IL-10 | TNFα | IL-6 | IL-8 | IL-10 | TNFα |
| Mothers | Genomes | **75.6%** | 64.0% | **70.6%** | **74.1%** | 6.9E-04 | 0.039 | 2.8E-03 | 2.8E-03 |
|  | Pathways | **72.1%** | 62.0% | 68.6% | 61.8% | 6.9E-04 | 0.067 | 5.6E-03 | 0.18 |
| Children | Genomes | **77.8%** | **76.0%** | **74.5%** | **75.0%** | 6.9E-04 | 6.9E-04 | 6.9E-04 | 2.8E-03 |
|  | Pathways | **75.6%** | 66.0% | 66.7% | 56.4% | 1.8E-03 | 0.02 | 0.01 | 0.45 |

**Table 5.**
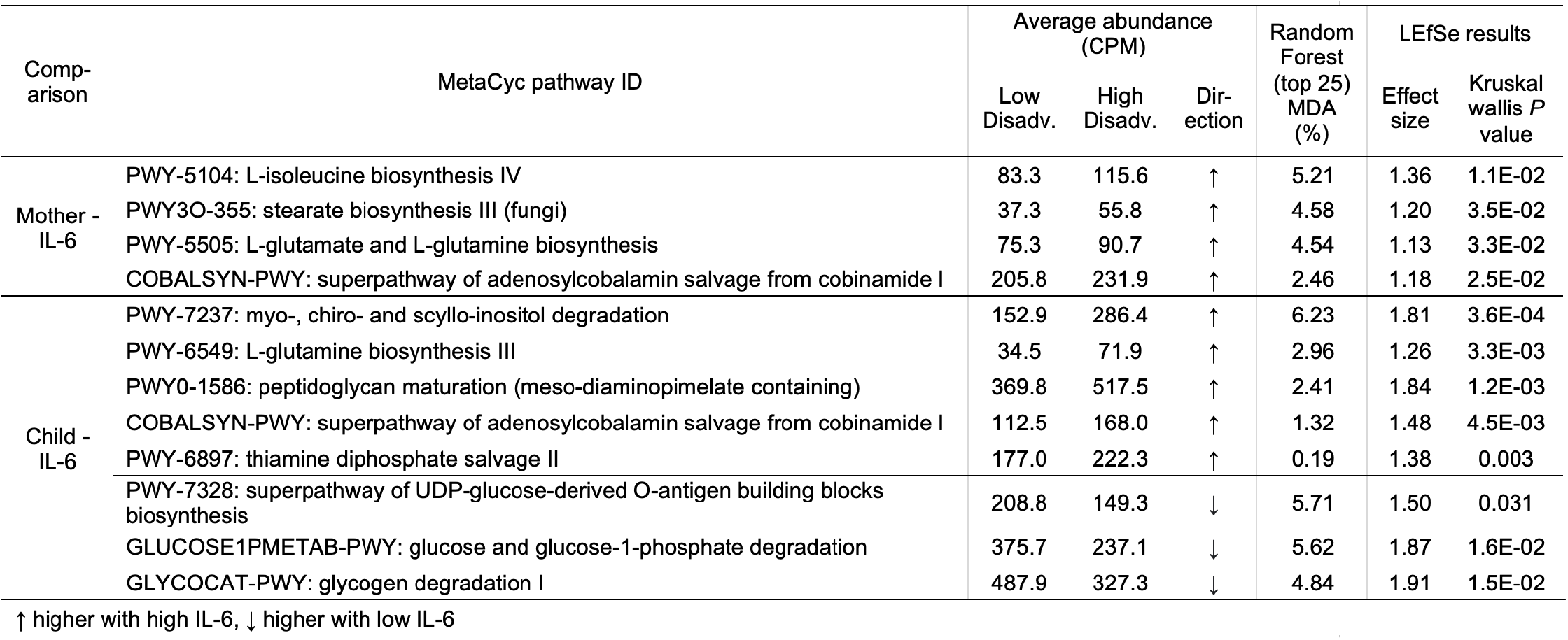
MetaCyc pathways differential abundance in the mothers and children, based on comparisons of high-vs-low IL-6 abundance. Based on MGS HUMAnN3 MetaCyc pathway abundance, RF can successfully classify mothers as low and high IL-6 73.3% of the time (*P* = 6.9×10^−4^ compared to random assignment; FDR-corrected binomial distribution test). Average abundance and association (high or low) are shown for the pathways predictive value in the top-25 RF model (ranked by mean decrease in accuracy of the RF model; MDA), and with LEfSe effect size ≥ 1. The -log of the Kruskal-Wallis test *P* values from LEfSe is also shown.

**Figure 8:**
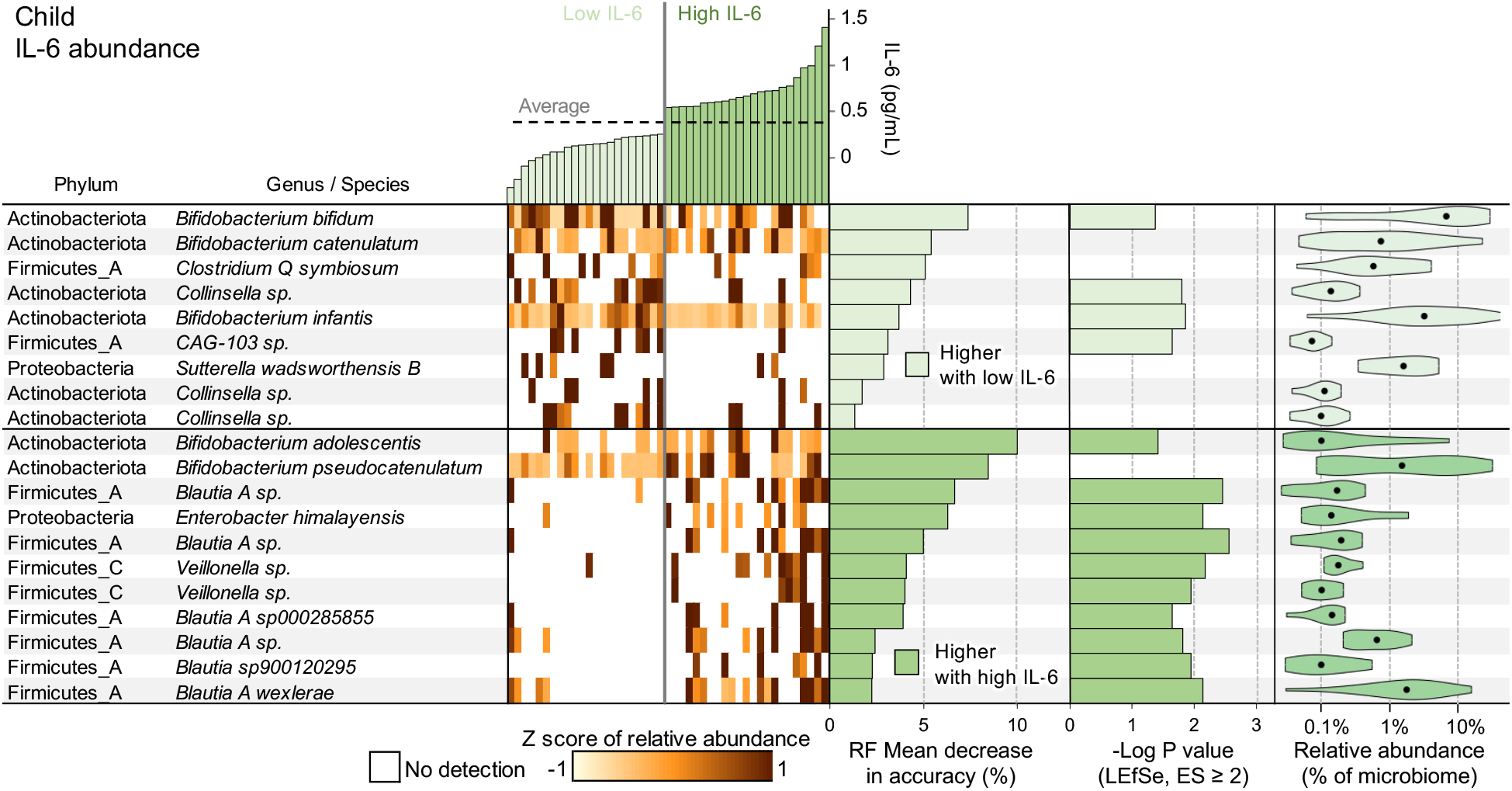
MGS genome differential abundance in children, based on comparisons of high-vs-low IL-6 abundance. Based on MGS genome profiles, Random Forest (RF) can successfully classify children into high and low IL-6 abundance 77.8% of the time (*P* = 6.0×10^−5^) compared to random assignment; FDR-corrected binomial distribution test). Taxonomy and relative abundance per sample for the genomes with the highest predictive value in the top-25 RF model are shown (ranked by mean decrease in accuracy of the RF model; MDA). Also displayed are -Log of the Kruskal-Wallis test P values from LEfSe (no value shown if the effect size was <2) and the overall abundance of the taxa when present.

## CONCLUSIONS

Our prospectively assembled cohort of mother-infant dyads, enabled us to quantify for the first time the impact of exposure to both, SD and PS on GM structure and function for mothers and their infants. Mothers and infants classified as “high’ (case) or “low” (control) SD/PS, show distinct discriminatory taxonomic, metabolic and inflammatory features that accurately ‘predict’ maternal prenatal SD and PS status over 80% of the time, with SD having greater predictive accuracy than PS. Mothers with high-SD/high-PS have highly variable microbiomes compared to low-SD/low PS mothers, reflecting greater permutations from environmental influences. The distinct nature of the taxonomic and functional GM predictors for SD compared to PS, indicate different underlying mechanisms driving the relationship.

The human GM modulates inflammatory cytokine production [8, 67] and has been linked to chronic inflammatory disorders [6]. We identified a significant relationship between the maternal prenatal and infant GM, and prenatal circulating cytokine concentrations in mothers. Prediction accuracy is highest and most consistent for IL-6 suggesting a contributing role to subtle chronic inflammation, although the discriminating taxa are distinct from SD. Elevated IL-6 concentrations have been linked to specific GM profiles in disease states among adults [68, 69] and recently to neuropathology when elevated during pregnancy in animal models [64, 65].

Our findings should be viewed within the strengths and limitations of the study. The finding are associations, and do not necessarily imply causality. Identification of strain level taxa and metabolic pathways through WMG, however, form the basis for testing mechanistic causation in preclinical models. Use of mother-infant dyads allowed interrogation of SD and PS between mother and infant GM, and examination of genomes and metabolic pathways. The population we studied is from a circumscribed region but contains a broad range of socio-economic backgrounds. We did not examine the role of race, because of the collinearity of race and SD, with no additional contribution of race in the model beyond that found with SD alone.

While longitudinal studies are needed to determine the stability of these GM signatures from early life to early childhood in children, the results identify unique features of the maternal and infant GMs and host response. Such findings should be pursued to better understand the effect of socioeconomic status and mental health determinants on GM health and stability. More critically, information on causal pathways triggered or sustained by the GM that affect child health and development could lead to new biomarkers and interventions. The potential malleability of the GM leaves room for optimism that unfavorable neurodevelopment outcomes might not be inevitable in children with living with high SD and PS values.

## METHODS

### Study Design and Cohort

The study population Consists of 121 mother-child dyads drawn from a larger parent study of 399 dyads, Early Life Adversity Biological Embedding and Risk for Developmental Precursors of Mental Disorders (eLABE; details provided in [29]). The eLABE study used a prospective observational design to examine the impact of pre- and post-natal psychological and social factors on infant neurodevelopment. Preganant women (N=395) and their offspring were recruited from the March of Dimes Prematurity Research Center at Washington University in St. Louis between 2017-2020, with delivery of singleton births at the Barnes Jewish Hospital in St. Louis [70]. This sub-cohort was chosen from the extremes of maternal social disadvantage and psychosocial stress, among women with relatively healthy offspring, who donated third trimester stools. Exclusions included multiple gestation, congenital malformations and infections, premature birth (< 37 weeks gestational age), maternal alcohol or drug use during pregnancy (excluding tobacco, marijuana, and maternal enteral steroid use). Race and ethnicity were based on maternal self reporting extracted from the medical record. Options included American Indian/Alaskan Native, Asian, Black or African American, Native Hawaiian/Pacific Islander, White, unknown, or other (free text) for race, and the following options for ethnicity: Hispanic/Latina, non-Hispanic/Latina, or unknown/not applicable for ethnicity. All procedures were approved by the Human Research Protection Office, informed consent was obtained from the mother for each dyad. The study was performed in accordance with Strengthening the Reporting of Observational Studies in Epidemiology (STROBE) guidelines [71].

### Maternal measures

At each trimester of pregnancy, measures of maternal depression, experiences of stress, as well as demographic information including insurance, education, address, and household composition were obtained from participants or extracted from the medical record by trained staff. Components of the two latent constructs, maternal social disadvantage and maternal psychosocial stress are previously described [29]. Briefly, the components of each latent variable included:

#### Maternal Social Disadvantage

<u>Insurance status</u> obtained was verified at the third trimseter from the medical record and maternal self reporting, <u>Income to Needs ratio</u> in each trimseter based on family income and houshold size (1.0 being the poverty line for the U.S.) was self reported, highest <u>maternal educational level was self reported</u>, the national <u>Area Deprivation Index</u> is a national multidimensional geotracking method based on census block data, providing percentile rankings of neighborhood disadvantage status [72], and maternal nutrition over the past year was categorized using the validated <u>Healthy Eating Index</u> (using National Cancer Institute. The Healthy Eating Index – Population Ratio Method. Updated December 14, 2021; [73]) obtained using the Diet History Questionnaire (DHQII).

#### Maternal Psychosocial Stress

In each trimester mothers completed the Edinburgh Postnatal Depression Scale (EPDS) [74], Perceived Stress scale (PSS) [75] at each trimester, averaged over trimesters a one-time lifetime STRAIN survey [76], a comprehensive measure of lifetime stressful and traumatic life events. Experiences of discrimination based on race were assessed using the Everyday Discrimination Scale [77].

#### Maternal medical risks

were defined by the Maternal Medical Risk score in pregnancy, a validated measure of maternal co-morbidities weighted by severity in pregnancy, and maternal pre-pregnancy Body Mass Index was extracted from the medical record.

### Infant measures

<u>Gestational age</u> was determined by the best obstetric estimate using last menstrual period or earliest ultrasound dating. <u>Birthweight and route of delivery</u> were extracted from the electronic medical record delivery note. <u>Breastfeeding data</u> was collected by parental reporting at the time of home stool sample collection and based on the Center for Disease Control Infant Feeding Practices II study food frequency checklist data [78, 79].

### Biological specimen collection and processing

Maternal blood samples for serum were collected from venous draws during routine clinic visits across each trimester, processed within 12 hours of collection, and stored at -80°C in 1 mL aliquots (details in [70]). Stools from mothers and infants were collected from home and processed as previously described [79]. Briefly maternal samples were collected during the third trimester, and infant stools were retrieved directly from the diaper and scooped into barcoded tubes. Stool samples were placed in insulated packaging (VWR) with U-tex gel packs (Fischer Scientific) and placed in home freezers. A community-based courier system available 24 hours per day was used to retrieve samples within 90 minutes. Weekend or overnight samples were taken to the courier office freezers (−20°C) and then delivered to the laboratory freezer (−80°C) during working hours; weekday stools were couriered directly to the laboratory. Previous testing showed no quality difference between temperatures used in delivery methods [80]. DNA was extracted from stool using the Qiagen (Hilden, Germany) QIAamp Power Fecal Pro DNA Mini Kit (50) and the automated QIAcube platform (Qiagen) as previously described [79, 81].

### Targeted GM profiling using V4-16S rRNA sequencing, data processing and analyses

DNA extracted from stool was sequenced on an Illumina MiSeq, producing 2×250bp paired-end reads spanning the V4 hypervariable region, for 242 samples (121 samples from mothers and 121 from their matched children). Data were imported into QIIME2 [82], using standard methods and the developer’s docker container (qiime2/core:2018.8). V4 region amplicons were assembled and denoised using the Qiime2 method ‘DADA2 denoise-paired’. Processed V4 amplicons were grouped into amplicon sequence variants (ASVs) with 100% sequence similarity. ASVs were classified using a pre-trained classifier based on SILVA (release 132) [83], a comprehensive database that provides accurate annotations [84]. ASV counts per sample were exported as biom files from a qiime2 artifact and converted into a human readable tsv file using “biom convert”. Read counts per sample were rarefied to 11,929 reads per sample (the lowest count among the 242 samples) using the “rrarefy” command in the R package “vegan” (version 2.5-7, https://CRAN.R-project.org/package=vegan), and normalized read counts were calculated per sample by dividing the number of reads associated with each ASV by the total number of reads assigned across ASVs. Taxonomic identifications used are directly provided by SILVA [83]. Raw 16S rRNA can be downloaded from public database (submission in progress).

### Whole Metagenome Shotgun (WMS) sequencing and GM profiling

Whole Metagenome Shotgun (WMS) datasets for 178 samples (89 samples from mothers and 89 from their respective children) were generated on the Illumina NovaSeq S4. For each sample ∼ 6Gbp was generated. The reads for all 178 samples were cleaned of barcodes, adapters and low-quality ends using Trimmomatic [85] (version 0.36). The BMTagger program (installed using conda, July 30th, 2020) was used to identify human contaminant reads using the human reference genome (GRCh38.98 [86]). Reads identified as human were removed to produce final paired-end fastq per each of the 178 samples. Raw WMS read data can be downloaded from public database (submission in progress).

The 178 WMS samples were mapped against the Unified Human Gastrointestinal Genome (UHGG) collection, comprising 204,938 nonredundant genomes from 4,644 gut prokaryotes, each theoretically representing an individual bacterial or archaeal species (95% average nucleotide identity [32]) using bowtie2 [39] (v2.3.5.1). The profile module of the inStrain [33] program was then run to generate sequencing breadth and depth of coverage statistics for every genome, in addition to nucleotide diversity measures per genome per sample. The depth of coverage values were normalized within every sample by dividing each genome’s depth by the sum of the depths across all genomes.

The 178 WMS samples were also used as input for HUMAnN [37] (version 3), which was ran from the biobakery/humann docker container (latest version as of October 2020) using the Chocophlan nucleotide database and Uniref90 [87] protein database. HUMAnN3 runs the MetaPhlAn [38] program as an intermediate step to assign organism-specific functional profiling, and we used the developer-provided Metaphlan3 [38] bowtie2 [39] database for this intermediate step. The HUMAnN3 pipeline was used to generate MetaCyc [36] pathway abundance per sample. The “humann_renorm_table” script (included in the HUMAnN3 distribution) was used to convert Reads Per Kilobase (RPK) values in the MetaCyc abundance table to a normalized value, Copies Per Million (CPM), which can be compared across samples.

### Statistical analysis

Significant differences between Low and High SD and PS sample sets based on metadata classifications (**Table 1, Supplementary Table S1**) were calculated using two-tailed T-tests with unequal variance for continuous variables and using Fischer exact tests for categorical data. Correction for multiple testing was not performed across tests, because significant differences indicate potential bias in the data. Significant differences in components of SD and PS were expected since samples were chosen from the extremes of phenotype for SD and PS to improve detection of GM differences between groups [29].

For the 16S/ASV sample analysis, Shannon index diversity values were calculated for each sample using the normalized read counts across all taxa using the “diversity” function in the “vegan” library in R, and Bray-Curtis distance diversity values were calculated using the “vegdist” function [88]. Correlation R^2^ values and Pearson r values were calculated using MS Excel, and the significance of the correlation was tested using the two-tailed *t* statistic with degrees of freedom N -1. ASV-based sample clustering was performed using the relative abundance profiles of all ASVs across all samples as input for Bray-Curtis dissimilarity-based clustering (complete linkage) using the “hclust” function in R, with additional metadata visualized using MS Excel. Significant differences in SD and PS between clusters were identified using ANOVA with a Post Hoc Tukey HSD test.

For the WMG sequence analysis, samples were divided into “high” and “low” SD and PS based on the distribution of these values across the sample set. Samples above the average value + 0.5 standard deviations were considered “high” and samples below the average value – 0.5 standard deviations were considered “low” (35 “low-SD”, 43 “high-SD, 36 “low-Psych” and 32 “high-Psych”; **Figure 1C**). The same approach was used to separate samples into “high” and “low” sample sets based on inflammatory marker data (IL-6, IL-8, IL-10 and TNFα).

To identify bacterial taxa that strongly predict mothers’ SD and PS scores, we analyzed taxonomic and pathway GM profiles using two approaches. First, a supervised machine-learning approach (Random Forest [40]) that identifies non-linear relationships from high dimensional and dependent data [40] and two-round approach using only the best 25 predictors (as previously described for gut microbiome associations with mental health measures [41]) was used to (i) quantify the ability to predict metadata classification based on the microbiome profiles, indicative of the overall association between the microbiome and the composite scores, and (ii) for each comparison, identify the specific genomes and pathways that most strongly differentiate between the high and low SD and PS scores. The generalization error of the model was evaluated by out of bag (OOB) error. The association of the metadata with the microbiome was quantified using the RF classification accuracy, and the significance of the accuracy was measured using FDR-corrected binomial distribution tests. RF model accuracy was also examined using receiver operating characteristic (ROC) curves, quantified using the area under the curve (AUC). Significance values for the ROC curves were assigned by Mann-Whitney U statistics [89], using the “roc.area” function in the R library “ROCR”.

Second, linear discriminant analysis effect size (LEfSe [42]), the most frequently used statistical tool to determine significant differences in microbiome member abundance [90], was used for differential genome abundance testing (default settings at an adjusted P ≤ 0.05 for significance) for the non-parametric factorial Kruskal-Wallis (KW) sum-rank test, and requiring a linear discriminant analysis (LDA) “effect size” of at least 2 in order to identify differentially abundant taxa). The same approach was used for the pathway analysis, but the “effect size” test cutoff applied was reduced to a value of 1 instead of 2, since the effect size is designed for the more sparse nature of metagenomic abundance data [42]. However, the same MDA and KW cutoffs were applied for pathway analysis.

## Data Availability

All data produced in the present work are contained in the manuscript.

## Acknowledgments

Funding, Washington University in St. Louis’ GTAC@MGI for library construction and sequencing, NIMH grant RO1 MH113883, (BBW, BAR, IMN, PPT, PJM, SKE, JLL, CER, TAS, CDS, DMB, GEM, EC, JM, MM) March of Dimes (SKE), Children’s Discovery Institute II MD-II-2015-489 (BBW, IMN), Biobank Core P30DK052574 (PIT).

## Author Contributions

Conceived and designed the study: BBW, MM JL, DB, CER CDS

Assembled the cohort, collected specimens: IMN, LL, SKE

Biological and clinical data database maintenance: IMN, REB, JDW, CH-M

Sample preparation and extraction of DNA: IMN, REB, JDW, CH-M

Performed data analysis: BAR, MM, JM, PJM, GEM,EC

Interpreted the data: BAR, MM, BBW, JM PPT, IMN

Wrote the paper: BAR, MM, BBW

All authors approve the manuscript for publication.

## Competing Interest

The authors have no competing interests to disclose.

## Additional information

We thank the families involved in the study, the MARCH of Dime Premature Research Center at Washington University, and the GTAC@MGI for the sequence data generation.

## Supplementary Figures

**Supplementary Figure S1:**
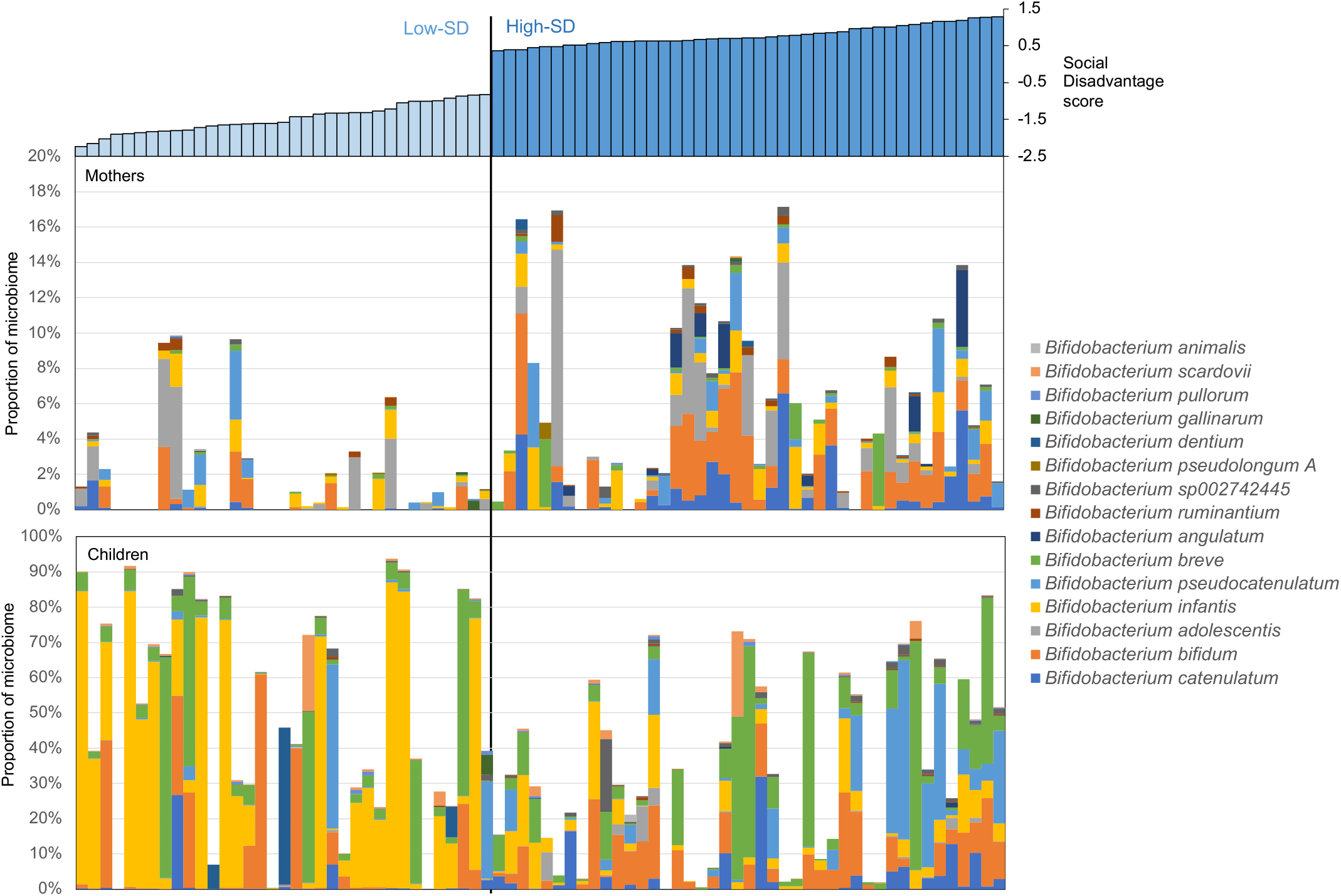
The relative abundance of *Bifidobacterium* species identified in all mothers and children in the high-SD vs low-SD comparisons.

**Supplementary Figure S2:**
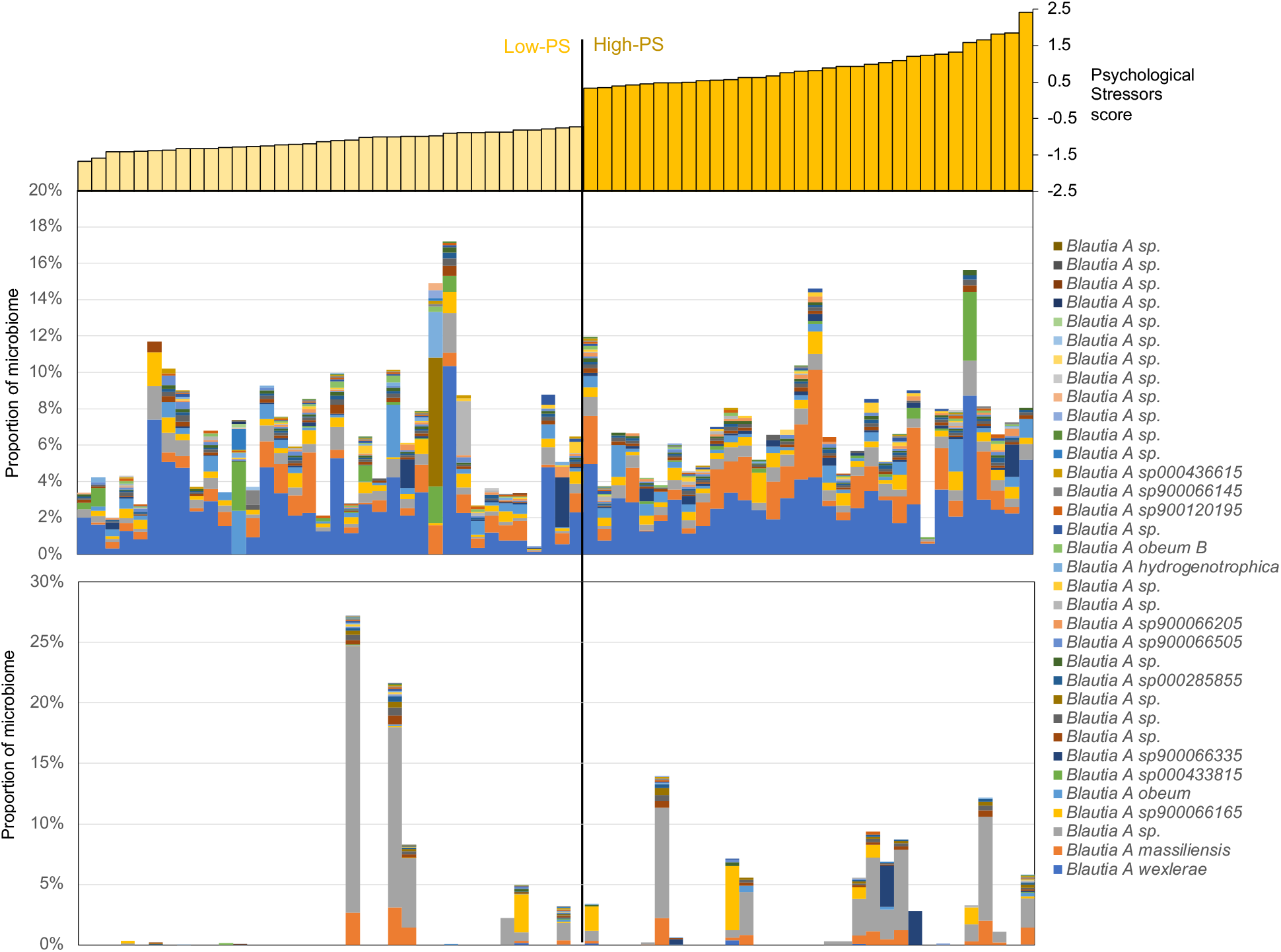
The relative abundance of *Blautia* species identified in all mothers and children in the high-SD vs low-SD comparisons.

**Supplementary Figure S3:**
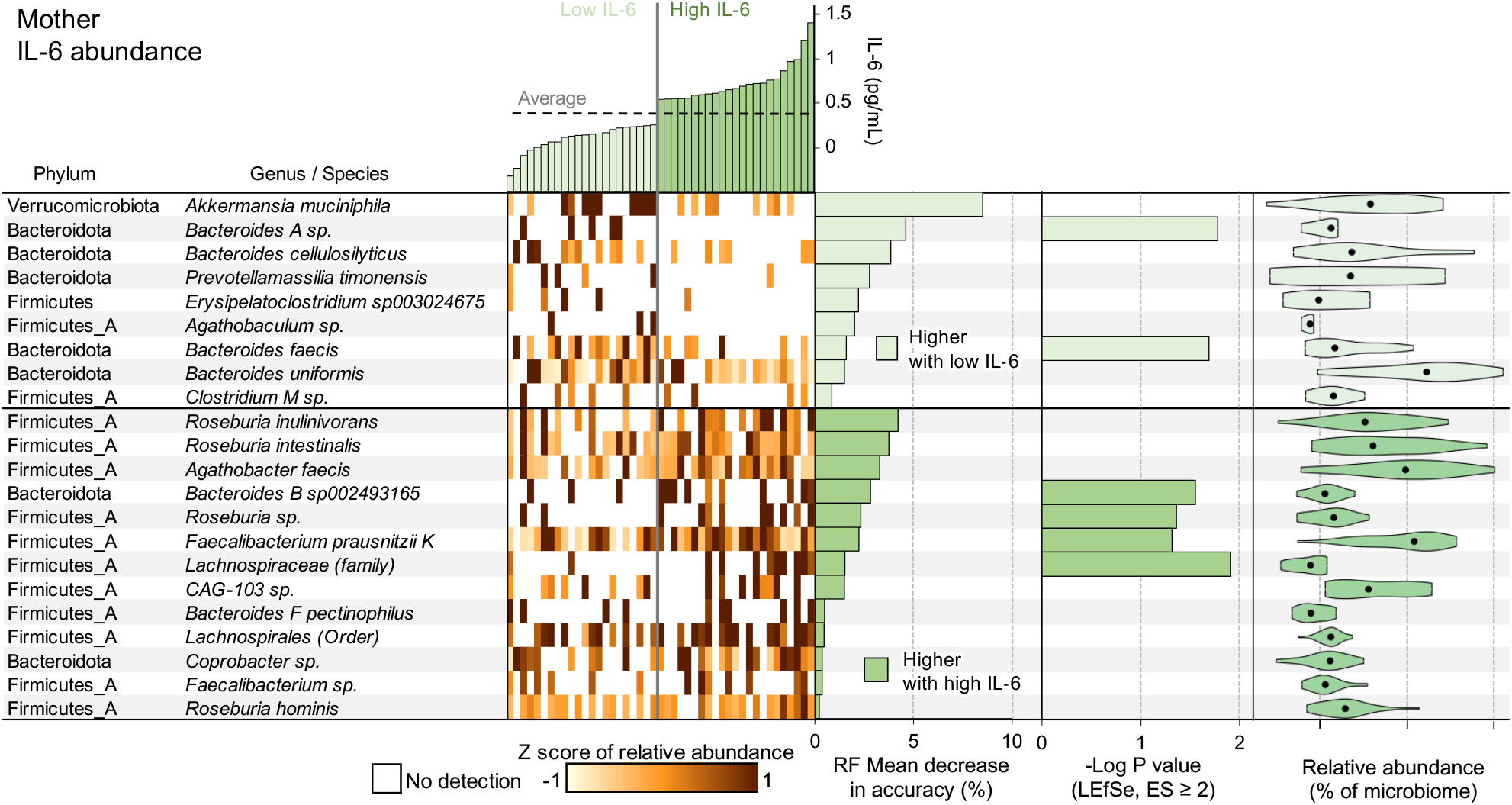
Based on WMS genome profiles, RF can successfully classify mothers as low and high IL-6 abundance 75.6% of the time (*P* = 2.2×10^−4^ compared to random assignment; FDR-corrected binomial distribution test). Taxonomy and relative abundance per sample for the genomes with the greatest predictive value in the top-25 RF model are shown (ranked by mean decrease in accuracy of the RF model; MDA). Also displayed are -Log of the Kruskal-Wallis test P values from LEfSe (no value shown if the effect size was <2) and the overall abundance of the taxa when present.

**Supplementary Figure S4:**
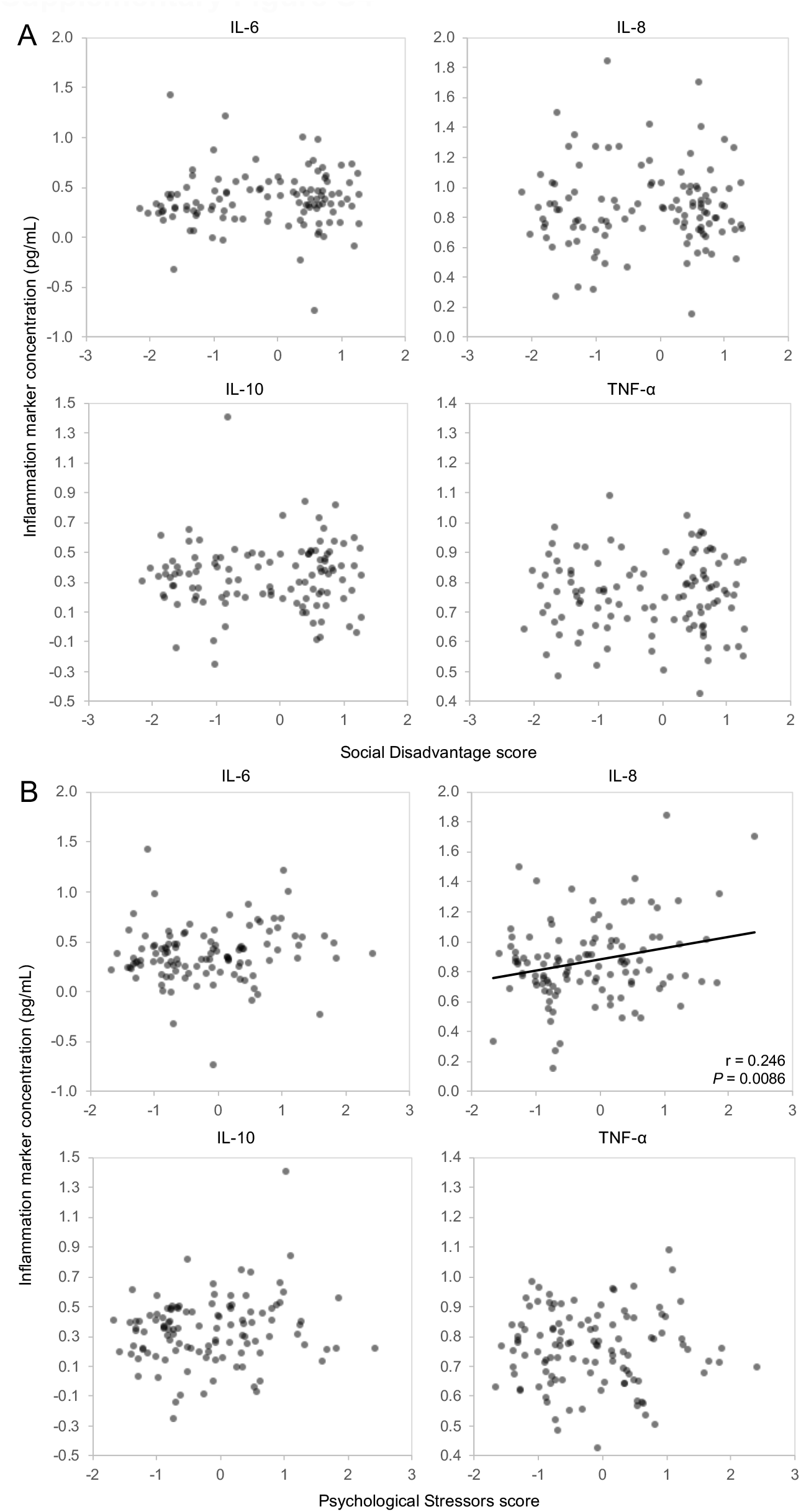
Comparison of Social Disadvantage scores and Psychological Stressors scores to maternal third trimester inflammatory marker serum concentrations. Only IL-8 and PS scores correlated significantly.

## Supplementary Tables

**Supplementary Table S1:** Complete patient characteristics at study entry for each of the primary comparisons of interest. “Low” and “High” Disadvantage and Psych scores are separated according to the distribution of the metadata, as shown in Figure 1.

**Supplementary Table S2:** Database of sample data, including metadata, indications of sample groups for each comparison, relative 16S ASV abundance, relative MGS genome abundance and relative MGS pathway abundance values.

**Supplementary Table S3**: Full ASV sequences for each unique ASV identifier from Table S2.

**Supplementary Table S4:** Differential abundance statistics for each UHGG genome in each comparison.

**Supplementary Table S5**: Differential abundance statistics for each MetaCyc pathway in each comparison.

## Notes

### Competing Interest Statement

The authors have declared no competing interest.

### Funding Statement

This study was funded by the Washington University in St. Louis GTAC@MGI for library construction and sequencing, NIMH grant RO1 MH113883, March of Dimes (SKE), Childrens Discovery Institute II MD-II-2015-489 (BBW, IMN), Biobank Core P30DK052574 (PIT).

### Author Declarations

The Human Research Protection Office of Washington University in St. Louis gave ethical approval for this work. Informed consent was obtained from the mother for each dyad. The study was performed in accordance with Strengthening the Reporting of Observational Studies in Epidemiology (STROBE) guidelines.

